# Modeling the effect of vaccination strategies in an Excel spreadsheet: The rate of vaccination, and not only the vaccination coverage, is a determinant for containing COVID-19 in urban areas

**DOI:** 10.1101/2021.01.06.21249365

**Authors:** Mario Moisés Alvarez, Sergio Bravo-González, Grissel Trujillo-de Santiago

## Abstract

We have investigated the importance of the rate of vaccination to contain COVID-19 in urban areas. We used an extremely simple epidemiological model that is amenable to implementation in an Excel spreadsheet and includes the demographics of social distancing, efficacy of massive testing and quarantine, and coverage and rate of vaccination as the main parameters to model the progression of COVID-19 pandemics in densely populated urban areas. Our model predicts that effective containment of pandemic progression in densely populated cities would be more effectively achieved by vaccination campaigns that consider the fast distribution and application of vaccines (i.e., 50% coverage in 6 months) while social distancing measures are still in place. Our results suggest that the rate of vaccination is more important than the overall vaccination coverage for containing COVID-19. In addition, our modeling indicates that widespread testing and quarantining of infected subjects would greatly benefit the success of vaccination campaigns. We envision this simple model as a friendly, readily accessible, and cost-effective tool for assisting health officials and local governments in the rational design/planning of vaccination strategies.

## Introduction

COVID-19 vaccines are finally in reach. In December 2020, the vaccines from Pfizer-BioNTech and Moderna-NIH were approved [1–3], after an unprecedentedly intense and rapid vaccine research crusade. Both of these vaccines are based on RNA technology [2,4–6] and exhibit efficacies of nearly 95%, as evaluated in clinical trials[7]. As vaccination campaigns have started already around the globe (e.g., in England, USA, Canada, and México), these and other vaccines are expected to obtain approval during January 2021 in different countries.

The availability of approved vaccines provides certainty in the international quest to contain the greatest pandemic that we have experience in modern times. However, the widespread deployment of vaccines still faces some challenges [8]. The extent and rate of vaccination in each country, and even in each city, will be determined by the global rate of vaccine manufacturing, the local logistics of distribution and application, and even public perception [9]. The cost of vaccines and vaccination campaigns is another aspect that will define the final availability (rate and coverage) of vaccines in a particular community. In the end, each country will have to design its own vaccination strategy based on cost, availability, public perceptions, and logistic considerations. A few economies will be able to implement fast and wide-coverage vaccination campaigns aimed at controlling COVID-19 progression within a few months. For most countries, vaccine campaigns will have to be sustained long-term efforts that extend throughout at least most of 2021 due to logistic limitations or cost (Moderna′s vaccine will cost ∼25 pounds dose^-1^; Pfizer′s vaccine will cost ∼15 pounds per dose) [7].

In this context, relevant questions related to the impact of different vaccination strategies remain unanswered. What fraction of the population do we have to cover in a vaccination campaign? How much does the rate of vaccination matter? After vaccination starts, how soon can we go back to our “normal” lives? Are social distancing and widespread testing still needed through the vaccination period?

Intuitively, we can infer the qualitative answer to some of these questions. After all, a vaccine is not effective immediately after application, and herd immunity will be only evident after a significant fraction of the population is immune. Both of the presently approved vaccines (Pfizer-BioNTech and Moderna-NIH) have a high effectiveness of η∼0.95 [7,10], but they require administration of two doses in a time frame of 21 or 30 days. Therefore, vaccinated subjects will most probably exhibit protective levels of anti-SARS-CoV-2 antibodies only at ∼30 to 45 days after they received the first dose. During that post-vaccination but pre-immune period, the pandemic will evolve, as a dynamic process driven by infective subjects (symptomatic and asymptomatic) that have not been diagnosed and quarantined and continue spreading the virus among the susceptible (not yet immune) population. Therefore, the interplay between vaccination and COVID-19 evolution in a particular community is a dynamic process, where several different rates compete.

The relevant rates are the local rate of infection, as influenced by the effectiveness of social distancing measures, the rate of testing and effective quarantining of infected subjects, and the rate of acquired immunity due to SARS-CoV-2 infection or vaccination. In turn, the rate of infection is affected by the effectiveness of all social distancing measures in that community. In principle, all these rates have to be considered when attempting to render an accurate forecast of the evolution of COVID-19 in a particular territory. Several studies [11,12] have used mathematical modeling to investigate the role of vaccine efficacy and vaccine coverage in the attenuation of the pandemic progression. However, one aspect that remains poorly explored is the assessment of the effect of different vaccination rates on the progression of COVID-19. Again, the effect of the rate of vaccination must be analyzed in the context of the dynamic process of infection and in consideration of the effect of the social distancing and testing efforts as well.

Recently, we introduced an epidemiological model formulation that explicitly considers demographic variables and epidemiological parameters to calculate the progression of COVID-19 in urban areas [13,14]. The model also considers the effectiveness of social distancing measures and of massive testing for expeditious identification and quarantining of infective subjects as inputs. In this communication, we have modified our original formulation to include the effect of vaccination coverage and rate, and we use this simple model to predict the pandemic progress in highly populated cities. We present a wide range of simulation scenarios for a hypothetical urban area, and we evaluate the relative effect of different schemes of vaccination (i.e., combinations of social distancing, testing intensity, and vaccination coverage and rate) on the evolution of COVID-19.

## Results and discussion

Here, we used a simple mathematical model (Figure 1) based on the use of two differential equations. 

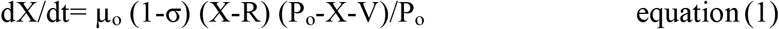

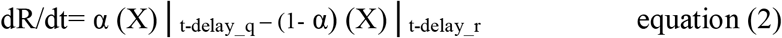

**Figure 1.**
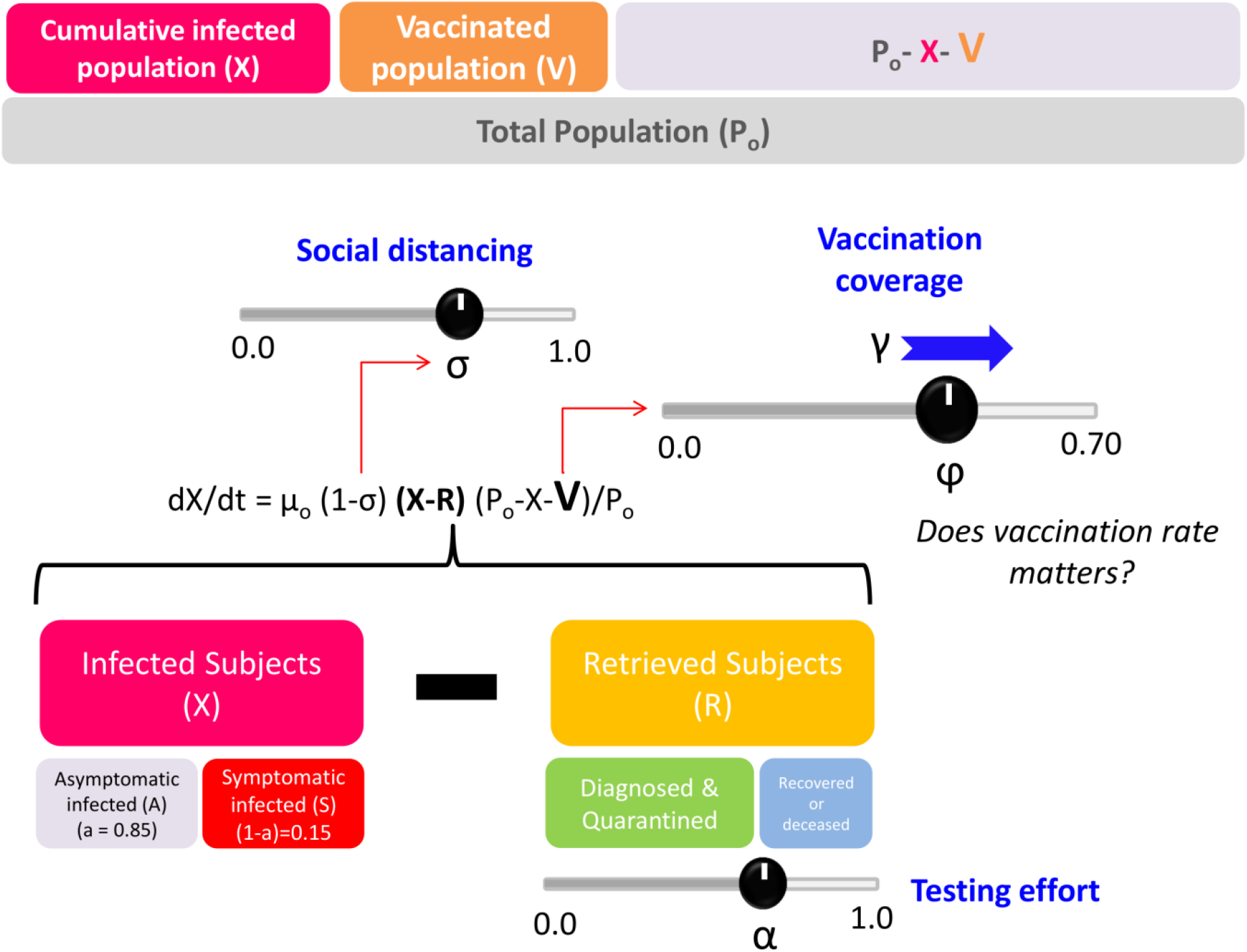
Schematic representation of a demographic mathematical model that considers social distancing (σ), intensity of the testing effort (α), vaccination coverage (φ), and vaccination rate (γ) to predict the evolution of pandemic COVID-19 in urban areas.

The first equation describes the rate of new infections (X), while the second accounts for the rate of accumulation of retrieved (R) subjects from the infective population, either because they have been effectively isolated form the rest of the population (i.e., hospitalized or quarantined after positive diagnosis). In this model, the quantity (X-R) is the driving force that sustains the local evolution of the pandemics and is proportional to the accumulation of new cases through a constant, namely the intrinsic rate of infection (µ_o_). However, this rate of generation of new cases is modulated by several factors, including the effective degree of activity in the region (1-σ). Here, α is a fraction that denotes the effective decrease in demographic density caused by the set of social distancing countermeasures (σ) currently established among that population (i.e., restrictions in social and economic activities, use of masks, restrictions in mobility, and all measures that contribute to an effective decrease in demographic density). The model also considers that the rate of generation of new cases is modulated by the fraction of the population that is susceptible to the infection (P_o_-X-V)/P_o_. This is precisely where the number of vaccinated subjects (V) plays a role by reducing the pool of susceptible individuals.

Using this simple epidemiological model, our aim was to reproduce representative settings for COVID-19 progression. For illustrative purposes, we have selected a representative urban area (i.e., more than 1,000,000 inhabitants and a population density above 4,000 habitants km^-2^). In all our simulations, we calculated the number of symptomatic subjects, the number of new cases per 1,000,000 of inhabitants, and the maximum hospital bed occupancy in that hypothetical region.

For all simulations presented here, we assumed that vaccination occurs immediately after the pandemic onset. Vaccination consists of the application of two vaccine dosages during 30 days. We assume that vaccinated subjects exhibit immunity only 45 days after receiving the first dose. Therefore, the effect of the vaccination campaign is only observable at day 45 after the pandemic onset in that locality. We also assume that social distancing is in effect during the entire vaccination campaign so that the level of activity in the city is decreased (and therefore the effective population density) by 50% (i.e., σ=0.50). Our simulations suggest that even aggressive vaccination campaigns (i.e., 70% vaccine coverage in 1.5 months) do not have substantial effects in the absence of social distancing and that they have only minor effects (Figure S1) even at moderate degrees of social distancing (i.e., σ=0.25).

We explored the range of vaccination coverage from 30 to 70%. Recent reports suggest that a vaccination coverage higher than 70% may be unattainable simply due to perceptions [15]. In surveys conducted in 28 nations, a pooled average of 20% of the participants expressed their intention to refuse SARS-CoV-2 vaccination. In addition, vaccine manufacturing is expected to be at a developing stage during 2021 [8]. Therefore, in all probability, the rate of manufacture of vaccines will be lower than their demand at least during the current year. In this context, reaching vaccine coverage higher than 70% seems challenging for most countries.

In a first set of simulations (Figure 2A and B), we investigated the effect of different rates of vaccination at fixed vaccination coverage under the following set of common assumptions: (a) the overall vaccination coverage will be limited to 30% of the population; (b) the intrinsic rate of infection is µ_o_ = 0.33, similar to actual values observed in densely populated urban areas, such as Mexico City and Madrid (Table S1); and (c) σ, the effective value of social distancing, is equal to 0.5. In addition, for these sets of simulations, the testing effort is such that only 15% of the infected subjects are tested and quarantined, while the rest of the infected subjects continue to be active until recovery. This strategy is consistent with that adopted by countries that have diagnosed essentially only those subjects who were symptomatic and asked for medical assistance (i.e., México, Chile, and Bolivia, with fewer than 2 tests per confirmed case).[16]

**Figure 2.**
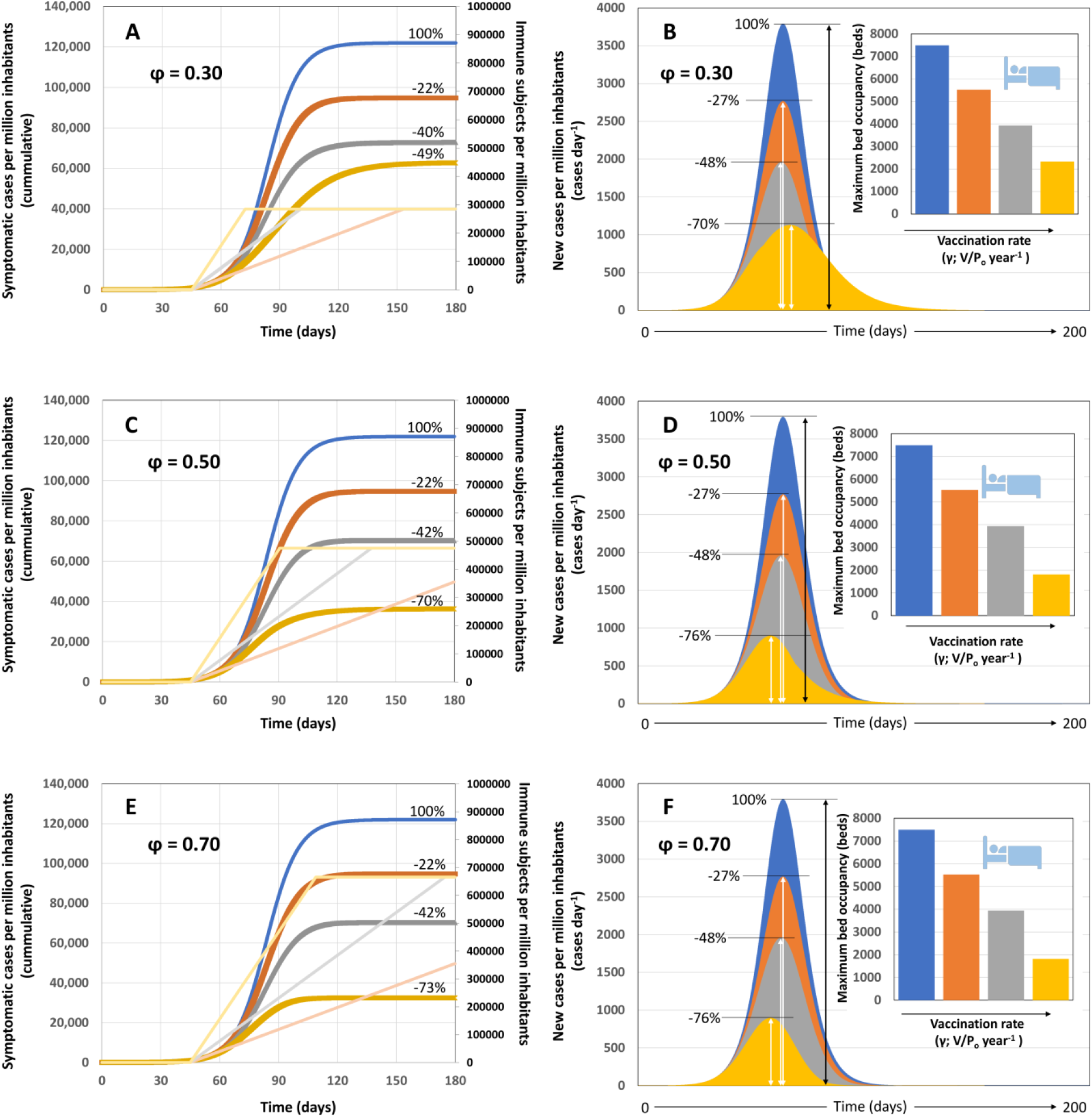
Scenarios of pandemic evolution for vaccination at different values of vaccination coverage and vaccination rates and under conditions of moderate social distancing (i.e., 50%) and basal values of testing effort (i.e.; α=0.15). (A) Cumulative number of infections, (B) daily number of new symptomatic cases and maximum bed occupancy (inset) for vaccination scenarios in which the vaccine coverage is kept constant at 30% P_o_ (φ=0.30), the effectiveness of the social distancing measures is 50% (σ=0.50), a basal level of testing is established (α=0.15), and different vaccination rates are established, such that: the entire population could be vaccinated within a year (vaccination at γ= P_o_ year^-1^; orange curve), within six months (vaccination at γ=2 P_o_ year^-1^; grey curve), or within three months (vaccination at γ=4 P_o_ year^-1^; yellow curve). A reference scenario without vaccination is included (blue line). Numbers indicate the percentage of reduction of symptomatic cases that results from the application of each vaccination rate with respect to the reference case. (C) Cumulative number of infections, (D) daily number of new symptomatic cases and maximum bed occupancy (inset) for vaccination scenarios in which the vaccine coverage is kept constant at 50% P_o_ (φ=0.50; σ=0.50; and α=0.15) and different vaccination rates are established such that: the entire population could be vaccinated within a year (vaccination at γ=P_o_ year^-1^; orange curve); within six months (vaccination at γ=2 P_o_ year^-1^; grey curve); or within three months (vaccination at γ=4 P_o_ year^-1^; yellow curve). (E) Cumulative number of infections, (F) daily number of new symptomatic cases and maximum bed occupancy (inset) for vaccination scenarios in which the vaccine coverage is kept constant at 70% P_o_ (φ=0.70; σ=0.50; and α=0.15) and different vaccination rates such that: the entire population could be vaccinated within a year (vaccination at γ=P_o_ year^-1^; orange curve); within six months (vaccination at γ=2 P_o_ year^-1^; grey curve); or within three months (vaccination at γ=4 P_o_ year^-1^; yellow curve). A reference scenario without vaccination is included in all panels (blue curve). Numbers indicate the percentage of reduction of symptomatic cases that results from the application of each vaccination rate with respect to the reference case. The number of immune subjects at different times, in accordance with different vaccination rates, is indicated with the same color code in the secondary axis.

Under this set of assumptions, we explored four different rates of vaccination (Figure 2A and B) to cover 30% of the population (φ=0.3) in 1 year (orange curves; γ= P_o_ year^-1^), in 6 months (grey curves; γ= 2 P_o_ year^-1^), and in 3 months (yellow curves; γ= 4 P_o_ year^-1^). These three rates of vaccination are equivalent to coverage of 100% P_o_ in one year (γ= P_o_ year^-^), six months (γ= 2 P_o_ year^-^), or three months (γ= 4 P_o_ year^-^).

The pandemic progression (number of cumulative symptomatic cases, and new infections per day) is indicated with blue curves for a reference scenario with a social distancing of 50% (i.e., an effective decrease of 50% in the demographic density) and a basal level of testing (i.e., only 15% of the infective subjects are tested and quarantined, while the rest of the infected subjects continue active until recovery). The results of our simulations show that increasing the rate of vaccination has a prominent effect on the epidemic curve.

Figure 2A shows the cumulative number of symptomatic cases at different vaccination rates when the vaccine coverage is limited to 30%. Figure 2B shows the new number of symptomatic cases per day at different vaccination rates and 30% coverage. The vaccination rate has a clear effect on both the shape and the area of the epidemic curves. Note that the peak of new infections also occurs sooner and is substantially lower as the vaccination rate is increased.

The total number of expected symptomatic cases (Figure 2A), the peak of new infections (Figure 2B), and the maximum bed occupation at hospitals (inset in Figure 2B) progressively and substantially decreased as the vaccination rate is increased. Evidently, a great benefit is attained by accelerating the distribution and application of vaccine as much as possible. The scenario of achieving a 30% coverage (300,000 vaccines per million of habitants) in three or six months seams feasible and reduces the overall number of symptomatic infected and the bed maximum bed occupancy in more than 50% with respect to the base case.

Figure 2C and D show the cumulative number of cases and the number of new infections per day, respectively, at different vaccination rates and at a vaccine coverage of 50% (φ=0.5). The maximum bed occupancy under this scenario is shown in the inset of Figure 2D. As before, we included a base case (blue curves) in which no vaccination occurs, whereas social distancing still has an effect equivalent to a decrease of 50% in the demographic density, and 15% of the infective subjects are diagnosed and quarantined. Similarly, Figure 2D and E show the cumulative number of cases and the number of new infections per day, respectively, at different vaccination rates and at a vaccine coverage of 70% (φ=0.7). The pandemic indicators (i.e., overall number of symptomatic subjects, peak of new cases per day and maximum bed occupancy) decrease modestly as vaccine coverage is increased from φ=0.30 to 0.50 or even 0.70. The benefit of increasing vaccine coverage at a given vaccination rate is substantially smaller than the benefit of increasing the rate of vaccination at a fixed vaccination coverage. Indeed, our results suggest that the overall vaccine coverage of 30% of the population at a rate of 100% P_o_ in 6 months is more effective than the vaccine coverage of 50% of the population at a rate of 100% P_o_ in one year, in terms of controlling the number of infections. Note that even increasing the coverage to 70% at the lowest rate does not provide the same benefits as a coverage of 50% achieved over a shorter period (e.g., 6 or 3 months). A modest coverage of 30% of the population at a rate of 100% P_o_ in 6 months provides better results than a higher coverage of 70% at a rate of 100% P_o_ in an entire year.

Figure 3 presents predictions for scenarios where 50% social distancing is implemented, but where the level of diagnostic testing is increased from a basal value (15% of infected are diagnosed and quarantined) to situations were 30% of infected patients (symptomatic and asymptomatic) are diagnosed within the first 5 days of viral shedding and are quarantined. This set of results allows observation of the effect of testing intensification combined with social distancing and vaccination. This would be a more realistic and recommended scenario. As before, the trends related to the reference scenario of 30% testing (α=0.30) and 50% social distance enforcement (σ = 0.50) are indicated in blue. The combination of moderate testing and vaccination renders better results than is achieved with basal testing and vaccination. The estimate of maximum bed occupancy is reduced by at least 50 % for all values of vaccination coverage and vaccination rates analyzed. For example, at the feasible scenario of 50% coverage at 100% P_o_ in 6 months, the maximum bed occupancy decreases from 5800 to 2030 beds if a moderate testing effort (α=0.30) is in place.

**Figure 3.**
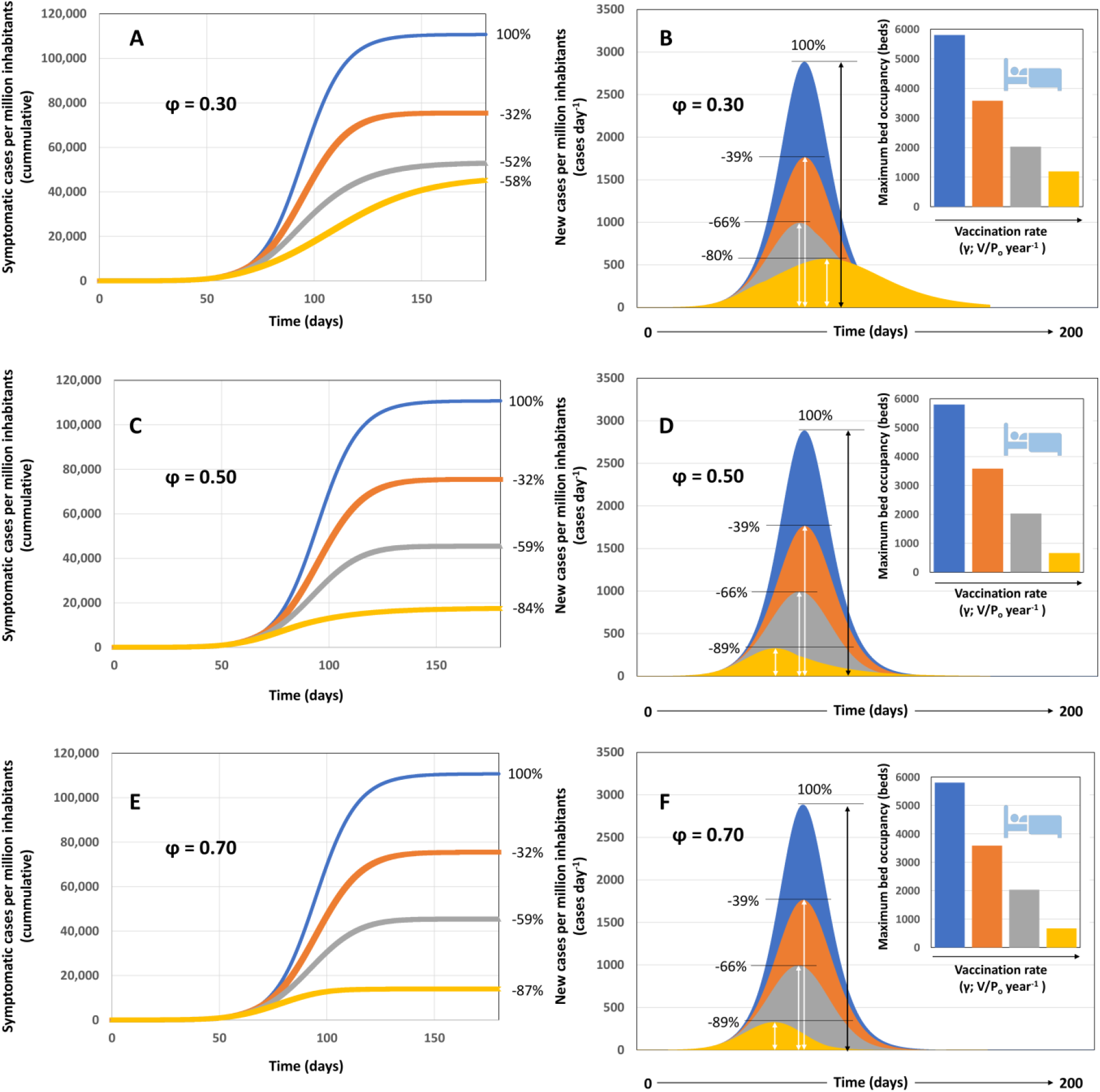
Scenarios of pandemic evolution for vaccination at different values of vaccination coverage and vaccination rates and under conditions of moderate social distancing (i.e., 50%) and enhanced testing effort (i.e., α=0.30). (A) Cumulative number of infections, (B) daily number of new symptomatic cases and maximum bed occupancy (inset) for vaccination scenarios in which the vaccine coverage is kept constant at 30% P_o_ (φ=0.30), the effectiveness of the social distancing measures is 50% (σ=0.50), a moderate level of testing is established (α=0.30), and different vaccination rates are established, such that: the entire population could be vaccinated within a year (vaccination at γ= P_o_ year^-1^; orange curve), within six months (vaccination at γ=2 P_o_ year^-1^; grey curve), or within three months (vaccination at γ=4 P_o_ year^-1^; yellow curve). A reference scenario without vaccination is included (blue line). Numbers indicate the percentage of reduction of symptomatic cases that results from the application of each vaccination rate with respect to the reference case. (C) Cumulative number of infections, (D) daily number of new symptomatic cases and maximum bed occupancy (inset) for vaccination scenarios in which the vaccine coverage is kept constant at 50% P_o_ (φ=0.50; σ=0.50; and α=0.30) and different vaccination rates are established such that: the entire population could be vaccinated within a year (vaccination at γ= P_o_ year^-1^; orange curve); within six months (vaccination at γ=2 P_o_ year^-1^; grey curve); or within three months (vaccination at γ=4 P_o_ year^-1^; yellow curve). (E) Cumulative number of infections, (F) daily number of new symptomatic cases and maximum bed occupancy (inset) for vaccination scenarios in which the vaccine coverage is kept constant at 70% P_o_ (φ=0.70; σ=0.50; and α=0.30) and different vaccination rates such that: the entire population could be vaccinated within a year (vaccination at γ= P_o_ year^-1^; orange curve); within six months (vaccination at γ=2 P_o_ year^-1^; grey curve); or within three months (vaccination at γ=4 P_o_ year^-1^; yellow curve). A reference scenario without vaccination is included in all panels (blue curve). Numbers indicate the percentage of reduction of symptomatic cases that results from the application of each vaccination rate with respect to the reference case. The number of immune subjects at different times, in accordance with different vaccination rates, is indicated with the same color code in the secondary axis.

As before, increasing the rate of vaccination is more efficacious than simply increasing vaccination coverage at a slow vaccination rate. For example, a vaccination strategy aimed at reaching 50% of the population at a rate of 100% P_o_ in 6 months results in the maximum bed occupancy of ∼2000 beds. By contrast, increasing the coverage to 70% at a rate of 100%P_o_ in one year yields a higher maximum bed occupancy (∼3600 beds).

A graphical summary of the results for different combinations of scenarios is presented in Figure 4. The effect of different vaccination rates is shown in terms of the number of symptomatic individuals at different vaccine coverage in (Figure 4A) and the maximum bed occupancy (Figure 4B). Lines drawn in different colors indicate the different values of vaccine coverage (i.e., 30, 50, and 70%). The vaccination rate exhibits a dominant effect on the parameters of the local evolution of pandemic COVID-19 and defines the number of symptomatic cases (Figure 4A) and the maximum bed occupancy (Figure 4B). Covering 70, 50, or 30% of the population at vaccination rates of 100% P_o_ in 12 or 6 months renders practically equivalent results. A higher vaccination coverage (50–70%) renders higher benefits than the lowest vaccination coverage (30%) only at the highest vaccination rate evaluated (i.e., vaccinating 100%P_o_ in 3 months).

**Figure 4.**
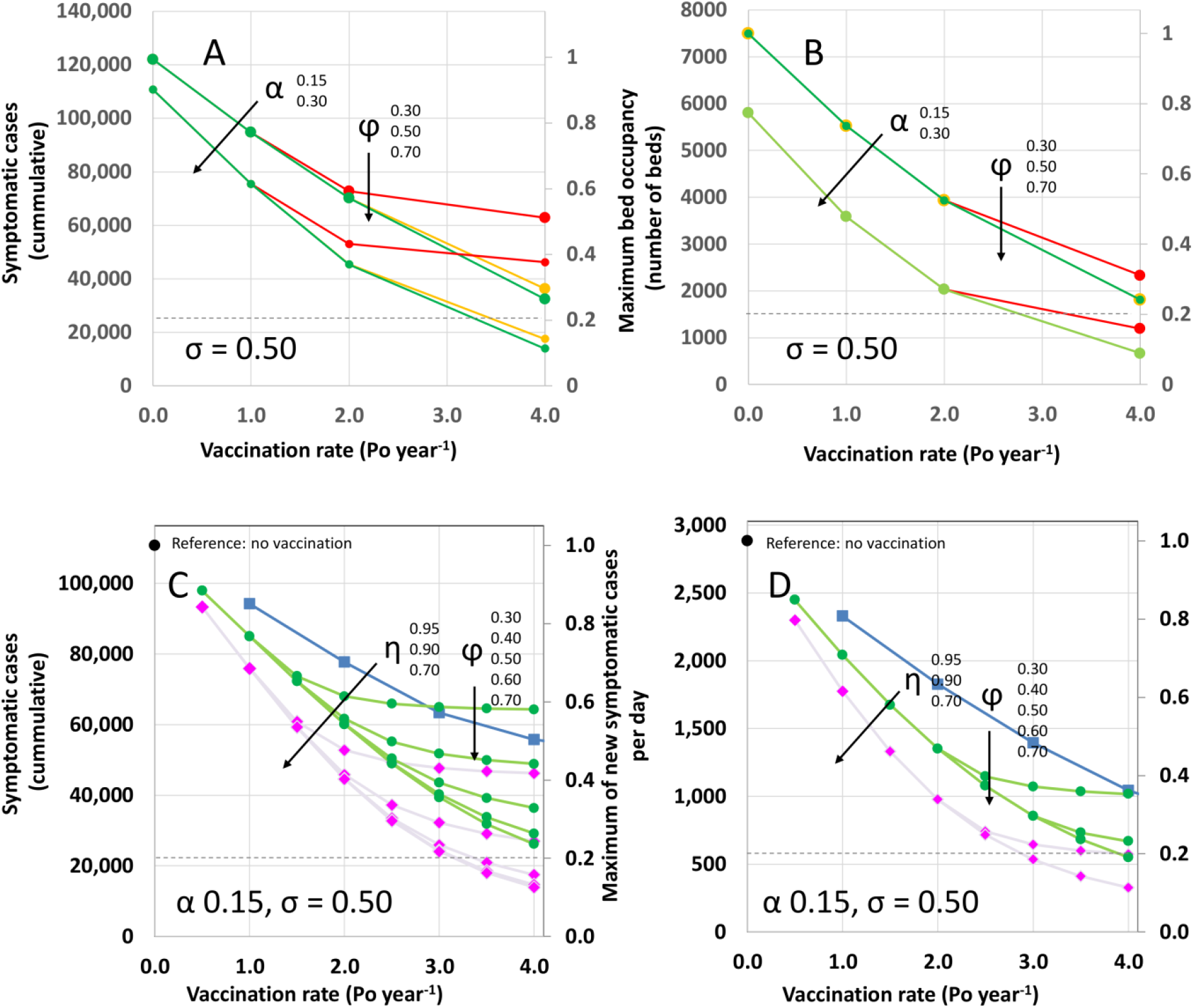
Effect of different values of vaccination coverage and vaccination rates on pandemic indicators. (A) Number of cumulative cases, and (B) maximum bed occupancy predicted in response to the application of different vaccination rates whereby the entire population could be vaccinated within a year (vaccination at γ=P_o_ year^-1^), within six months (vaccination at γ=2 P_o_ year^-1^), or within three months (vaccination at γ=4 P_o_ year^-1^) at different values of vaccine coverage: 30% P_o_ (ϕ= 0.30: red), 50% P_o_ (ϕ= 0.50: yellow), and 70% P_o_ (ϕ= 0.30: green). Curves for two different values of the coefficient of testing effort are presented (α=0.15 and α=0.30). The secondary axis indicates fractional values with respect to the same reference (i.e., no vaccination, moderate social distancing (σ=0. 50), and basal testing effort (α=0. 15). (C) Number of cumulative cases, and (D) maximum number of new symptomatic cases per day predicted in response to the application of different vaccination rates (γ= 0.5 P_o_ to 4.0 year^-1^) and at different values of vaccine coverage (from 30% to 70%; ϕ= 0.30 to 0.70). Curves for three different values of vaccination efficacies are presented (η=0.95; magenta rhombs, η=0.90; blue squares, and η=0.70; green circles). The secondary axis indicates fractional values with respect to the same reference (i.e., no vaccination, moderate social distancing (σ=0. 50), and moderate testing effort (α=0. 30).

In Figure 4A,B we selected a reduced set of scenarios to illustrate relevant relationships between vaccination rate and coverage and reduction in some of the indicators of the local evolution of pandemic COVID-19. However, the demographic model that we used is flexible enough to simulate a wide range of additional scenarios that combine social distancing, testing effort, vaccination coverage and rate, and vaccination efficacy (Table S2). In Table S2, five different indicators are calculated for different vaccination scenarios, including the day of the epidemic peak, the number of new infection cases at the epidemic peak, the cumulative number of symptomatic infections after 150 days of the local pandemic onset, the maximum bed occupancy, and the number of fatalities at different case fatality rates. Regardless of vaccination coverage, a fast vaccination rate (i.e., equal than or higher than 2 P_o_) renders reductions of more than 60% of symptomatic cases and maximum bed occupancies.

For example, Figure 4C and D present an analysis of the effect of the use of vaccines with different efficacies (i.e., η = 0.95, 0.90, and 0.70). These type of analysis may be highly useful for this and other epidemiology emergencies since constrains in the availability, cost, or logistics of vaccine storage or distribution will required countries to fulfill their vaccination requirements using not one but several vaccine alternatives.

## Conclusions

Within the next weeks, each county will have to design its own vaccination strategy based on cost, availability, public perceptions, and logistic considerations. Here, we aim to provide a friendly, easy-to-use modeling tool to assist governments and local health officials to rationally design vaccination campaigns.

We constructed a simple demographic model based on two differential equations that follow the accumulation of the number of infected subjects and the number of subjects retrieved from the infective population. This model formulation is extremely flexible and enables the prediction of the evolution of pandemic COVID-19 in urban areas based on simple reported parameters related to COVID-19 epidemiology and under different scenarios that may consider different degrees of social distancing and/or intensities of the testing effort, and different vaccination strategies. In this study, we focus on analyzing the effect of different levels of vaccination coverage and different vaccination rates on the local progression of COVID-19 in an urban area.

Our results suggest that the rate of vaccination is highly relevant in controlling the evolution of COVID-19 in highly populated cities. Based on the predictions of our model, we recommend the implementation of vaccination campaigns capable of achieving an overall coverage of at least 50% in short time periods (ideally within 3 or 6 months). In general, a faster deployment of vaccines should be preferred over a higher vaccine coverage that implies slower vaccination rates. Our model predicts that a vaccination campaign covering only 50% of the population in 3 or 6 months is more effective in controlling COVID-19 progression than is a vaccination strategy with a final coverage of 70% in longer times.

Importantly, our results clearly show that social distancing measures, including the use of face masks and restriction of regular social and economic activities, must not be lifted during at least the first three months of any vaccination campaigns. Only the combination of social distancing and vaccination enables the control of COVID-19 progression in highly populated cities within a time frame of 3–4 months. In addition, the intensification of the testing effort during vaccination yields relevant reductions in terms of decreasing maximum bed occupancy and the number of symptomatic cases through the implementation of vaccination campaigns.

## Materials and methods

### Mathematical model

We used an epidemiological model for the propagation of COVID-19 in urban areas that considers two variable populations of individuals, infected (X) and retrieved (R) (Figure 1). The cumulative number of infected patients (X) is the total number of subjects among the population that have been infected by SARS-CoV-2. The number of retrieved subjects is defined as the number of inhabitants that have been retrieved from the general population and therefore are not contributing to the propagation of COVID-19. Retrieved subjects include those who have recovered from the infection and do not shed virus, quarantined individuals, and deceased patients. The model assumes that infection results in short-term immunity upon recovery (at least 12 months). This assumption is based in experimental evidence that suggests that rhesus macaques that recovered from SARS-CoV-2 infection could not be reinfected [17]. However, the acquisition of full immunity to reinfection has not been confirmed in humans, although it is well documented for other coronavirus infections, such as SARS and MERS[18,19].

The interplay between these two main populations (X and R) and other subpopulations that include asymptomatic infected (A), symptomatic infected (S), and deceased (D) is determined by a set of demographic and clinical/epidemiological parameters (Table 1). The clinical parameters include an intrinsic infection rate constant (µ_o_) that can be calculated from the initial stage of the pandemic in that particular region, the fraction of asymptomatic patients (a), the delay between the period of viral shedding by an infected patient (delay_r), the period from the onset of shedding to the result of first diagnosis and quarantine in the fraction of patients effectively diagnosed (delay_q), and the fraction of infected patients effectively diagnosed and retrieved from the population (α). Demographic parameters include the population of the region (P_o_), the effect of social distancing (σ) in terms of effectively decreasing the demographic density, the fraction of infected individuals retrieved from the population due to massive and effective testing (α), and the vaccine coverage (φ) and vaccination rate (γ).

The model is based on a set of two simple differential equations. 

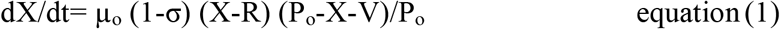

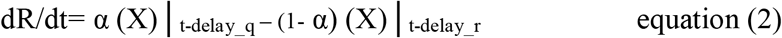

The first equation of the set (equation 1) states that the rate of accumulation of infected habitants (asymptomatic and symptomatic) in an urban area (assumed to be a closed system) is proportional to the number of infective subjects present in that population at a given point (X-R) and the fraction of the population still susceptible to infection ((P_o_-X-V)/P_o_). Note that the number of infective subjects is given by the difference between the accumulated number of infected subjects (X) and the number of retrieved subjects (R). The fraction of the susceptible population decreases over time, as more inhabitants in the community get infected (X) or are vaccinated (V). The proportionality constant in equation 1 (µ_o_) is an intrinsic rate of infection that is weighted by the effective fractional reduction of social distancing on the population density (1-σ).

The second equation (equation 2) describes the rate at which infected patients are retrieved from the infective population. Eventually, all infected subjects are retrieved from the population of infected individuals, but this occurs at distinct rates. A fraction of infected individuals (α) is effectively retrieved from the general population soon after the onset of symptoms or after a positive diagnosis. Another fraction of infected subjects (1-α) is not effectively retrieved from the population until they have recovered or died from the disease. Therefore, in our formulation, the overall rate of retrieval (dR/dt) has two distinctive contributions, each one associated with different terms on the right-hand side of equation 2. The first term accounts for the active rate of retrieving infected patients through the diagnosis and quarantine of SARS-CoV-2 positive subjects. For this term, the delay from the onset of virus shedding to positive diagnosis and quarantine (delay_q) is considered short (*i.e*., between two to five days), to account for a reasonable time between the positive diagnosis and the action of quarantine. In our model formulation, this term is represented by (1-α). A second term relates to the recovery or death of infected patients (symptomatic or asymptomatic) and is represented by the integral of all infected subjects recovered or deceased from the onset of the epidemic episode in the region, considering a delay of 21 days (delay_r), which accounts for the average time of recovery of an infected individual. Note that the simultaneous solution of equations 1 and 2 is sufficient to describe the evolution of the number of asymptomatic individuals (A), symptomatic individuals (S), and deceased patients (D) through the specification of several constants and simple relations. 

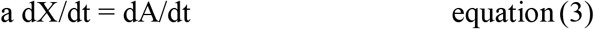

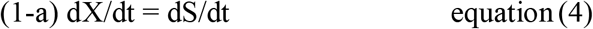

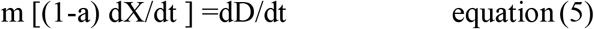

Here, **a** is the fraction of asymptomatic subjects among the infected population, (1-**a**) is the fraction of infected individuals that exhibit symptoms, and **m** is the mortality rate expressed as a fraction of symptomatic individuals.

This model relies on some basic assumptions that are sustained in clinical or epidemiological data.

### Parameter values and assumptions

The fraction of asymptomatic infected (a) is one of the critical inputs for the model; it determines the final and maximum feasible threshold of symptomatic infected. The current evidence is not yet sufficient to support a conclusive value for this parameter. For the simulations presented here, we set a=0.85, based on a recent serological study conducted in New York City (NYC) that found anti-SARS-CoV-2 IGGs among 21.2% of the population [20]. This serological result, combined with simulation work, suggests that nearly 85% of exposed New Yorkers were asymptomatic or exhibited minor symptoms. In addition, the average time of sickness was set at 21 days in our simulations, as this is within the reported range of 14 to 32 days[21], with a median time to recovery of 21 days[22]. Viral shedding can last for three to four weeks after the onset of symptoms, with a peak at day 10-11.[23] Therefore, we assume that all those infected not quarantined could continue to transmit the virus until full recovery (21 days). Similarly, asymptomatic patients are only removed from the pool of susceptible persons after full recovery. Asymptomatic patients are considered part of the population capable of transmitting COVID-19; reported evidence that suggests that asymptomatic subjects (or minimally symptomatic patients) may exhibit similar viral loads [24] to those of symptomatic patients and may be active transmitters of the disease [25,26].

The average fraction of deceased patients worldwide is estimated as 0.029 of those infected 21 days before [27]. We also consider that the average time for bed occupancy of hospitalized patients is 14 days. The estimated average hospitalization stays range from 9.3 to 13 days in the United States [28] and China [29,30], but much longer stays were reported in intensive care units in Italy (20 to 25 days)[31]. Anecdotal data collected in México suggests that hospitalization stays of at least two weeks are a more accurate figure for Latin American societies.

For illustrative purposes, we set the value of µ_o_ (the intrinsic rate of infection) in 0.33 day^-1^. This value is similar to that observed in Sao Paulo (∼0.308), Mexico City (∼0.329), Toronto (∼0.330), Lisbon (∼0.341), Madrid (∼0.358) and London (0.362), at the local onset of pandemic COVID-19 in 2020. Other urban areas have exhibited values of µ_o_ between 0.2 and 0.65 day^-1^ (Table S1). The value of µ_o_ (i.e., the intrinsic rate of infectivity of SARS-CoV2 before interventions) can be calculated for any urban area assuming that the initial rate of propagation is d(X)/dt = µ_o_ [X], where [X] is the number of initially infected subjects. Subsequently, the intrinsic rate of infection can be calculated from the initial slope of a plot of ln [X] *vs* time, which is a usual procedure for the calculation of intrinsic growth rates in cell culture scenarios under the assumption of a first-order rate growth dependence.

## Data Availability

All data related to this manuscript has been made available within the manuscript (as references) or as supplementary material.
The model used to generate the data discussed in this manuscript is also included as supplementary material.

## Acknowledgments

MMA, SBG and GTdS acknowledge the funding received from CONACyT (Consejo Nacional de Ciencia y Tecnología, México) and Tecnológico de Monterrey.

## Author contributions

MMA and GTdS collected and analyzed the epidemiology data. MMA formulated the model. MMA and SBG ran the simulations. MMA and GTdS analyzed and interpreted the simulation results and wrote the manuscript. All authors reviewed and approved the manuscript.

## Supplementary information

**Figure S1.**
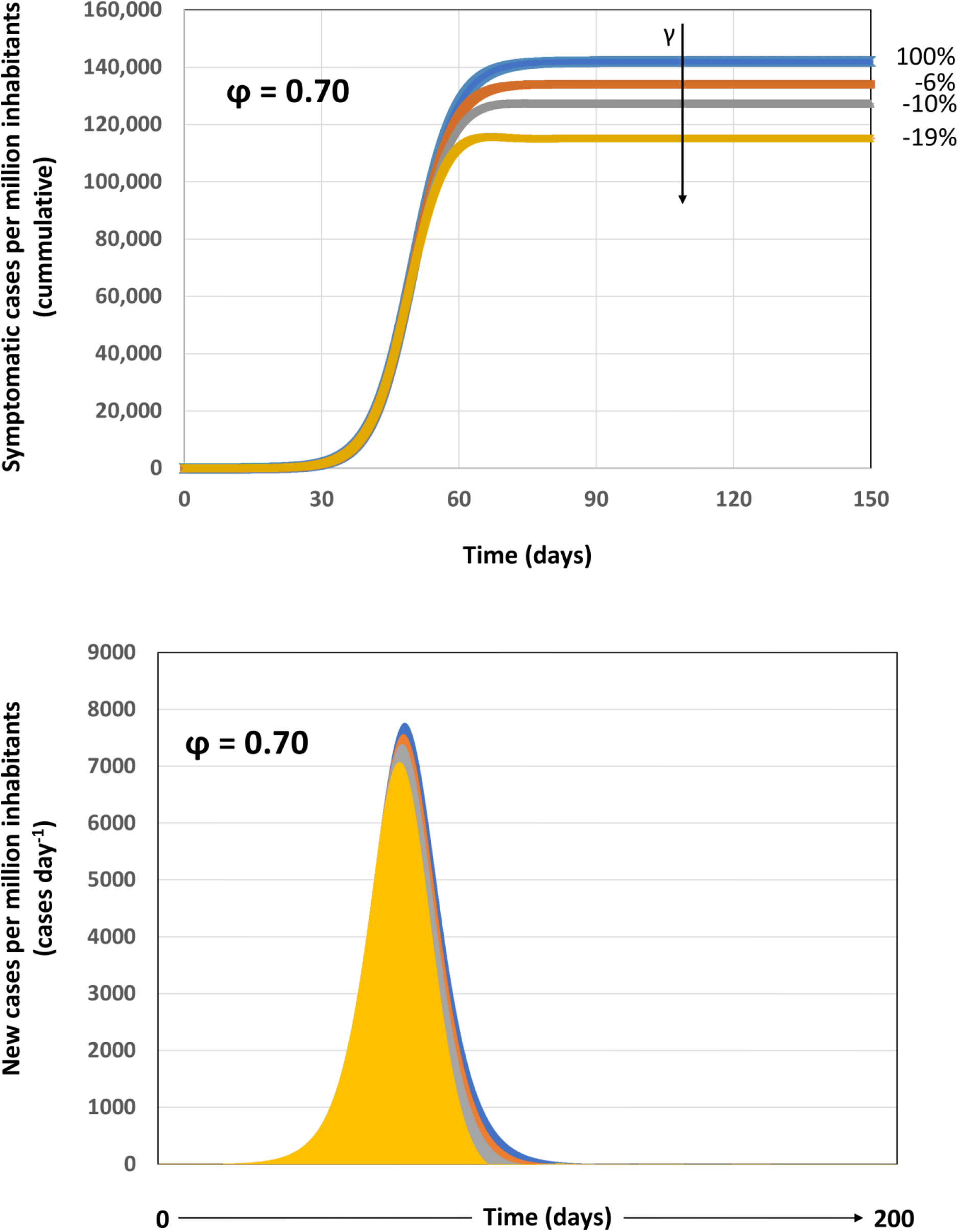
**Scenarios of pandemic evolution during the vaccination of 70% of the population at different vaccination rates under conditions of modest social distancing (i.e., 25%)**. (A) The cumulative number of infections, (B) the number of new infections per day and the maximum bed occupancy (inset) are presented for vaccination scenarios in which the vaccine coverage is kept constant at 70% P_o_ (φ=0.70), the effectiveness of the social distancing measures is 20% (σ=0.20), a basal level of testing is established (α=0.15), and different vaccination rates are imposed, such that: the entire population could be vaccinated within a year (vaccination at γ=0.0027 P_o_ day^-1^; orange curve), within six months (vaccination at γ=0.0054 P_o_ day^-1^; grey curve), or within three months (vaccination at γ=0.0108 P_o_ day^-1^; yellow curve). A reference scenario without vaccination is included (blue line). Numbers indicate the percentage of reduction of symptomatic cases that results from the application of each vaccination rate with respect to the reference case.

**Table S1.**
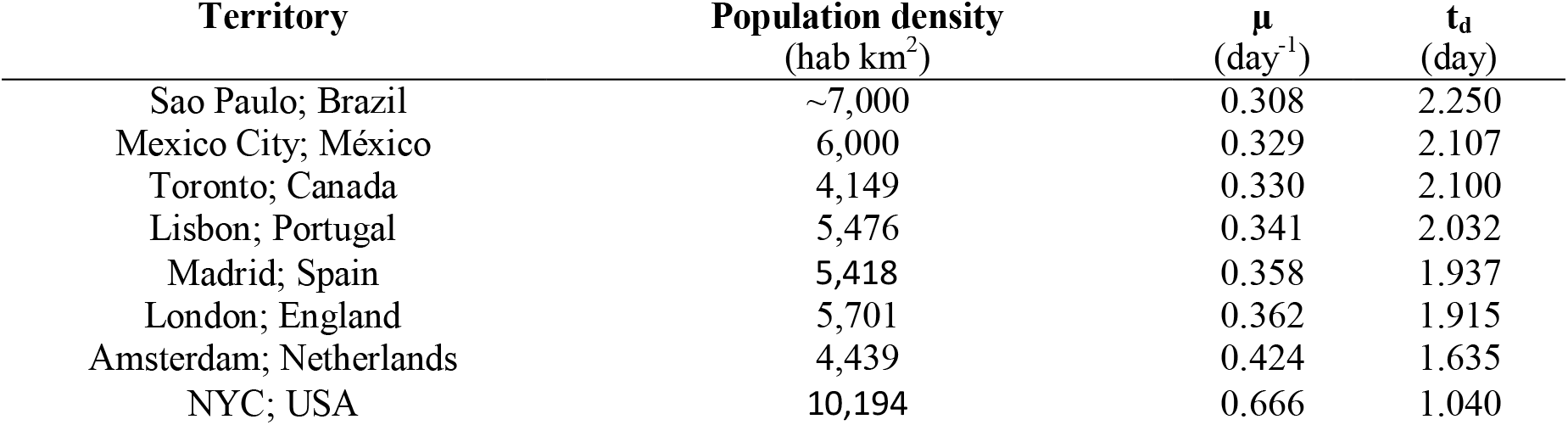
Specific infection rates (µ_o_) and the associated doubling times (t_d_) for COVID-19 infection in different geographic regions. Note that the intrinsic rate of infection ranges between 0.30 and 0.37 for territories with population densities between 4,000 and 7,000 hab km^2^.

**Table S2.**
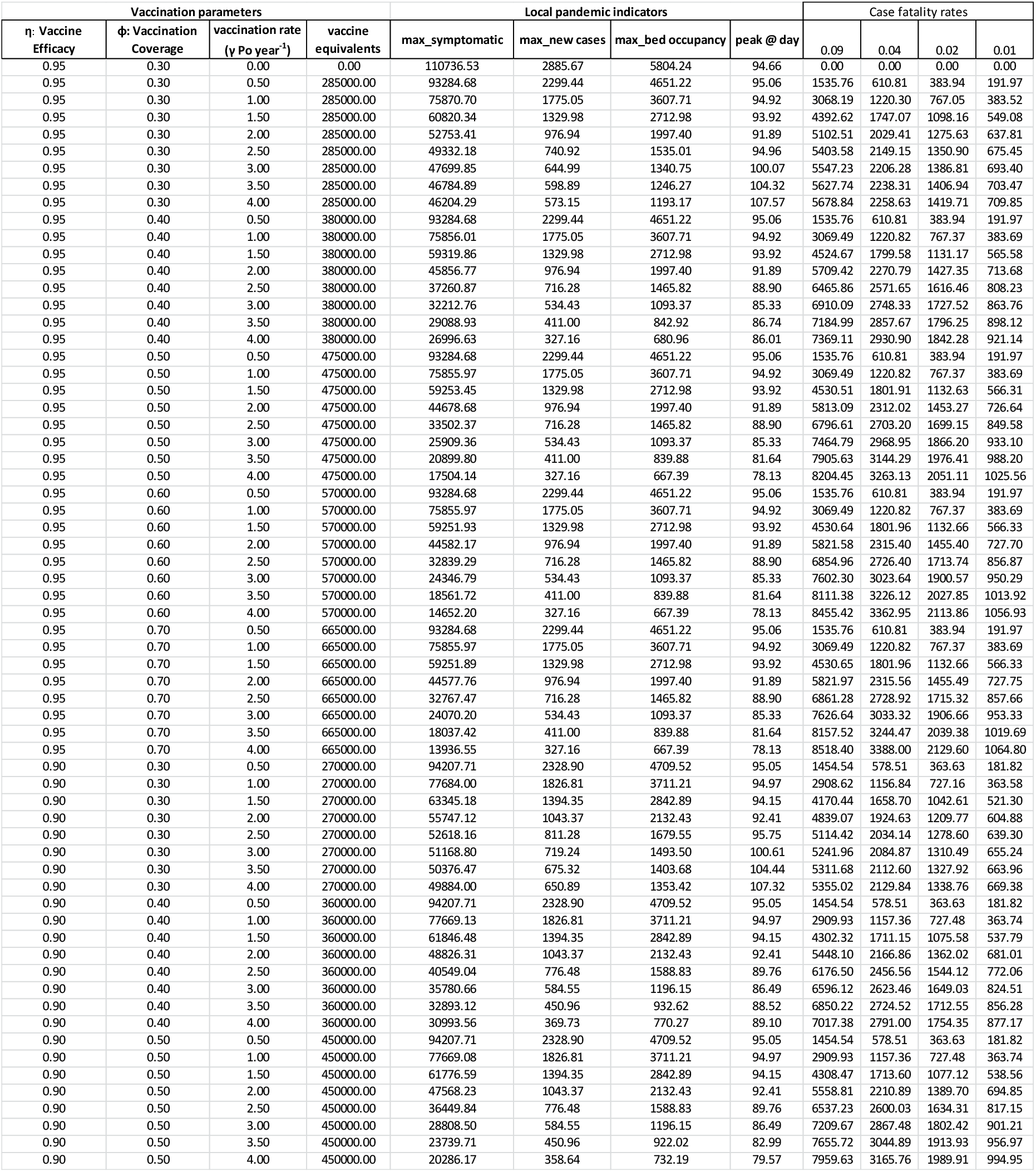
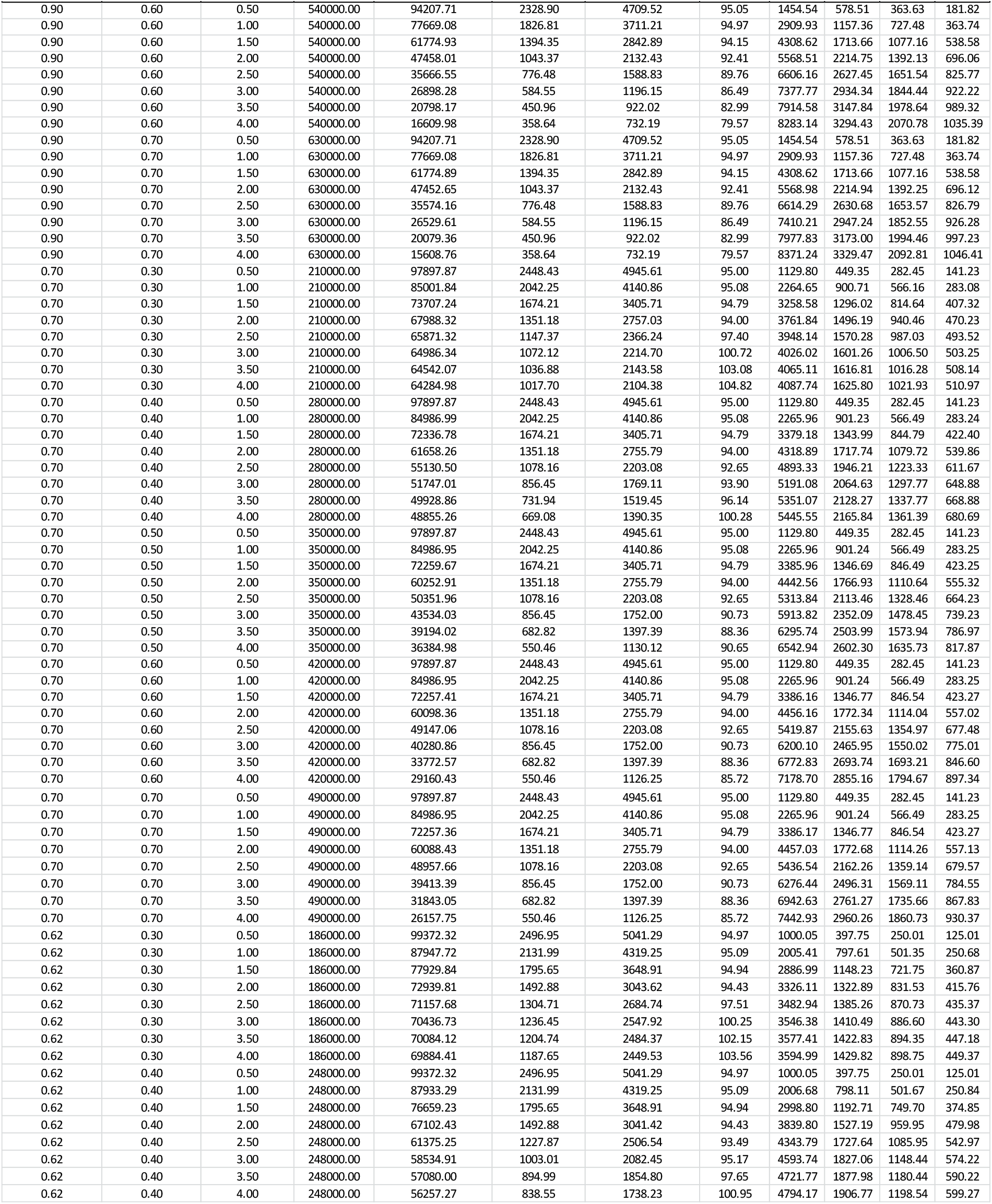
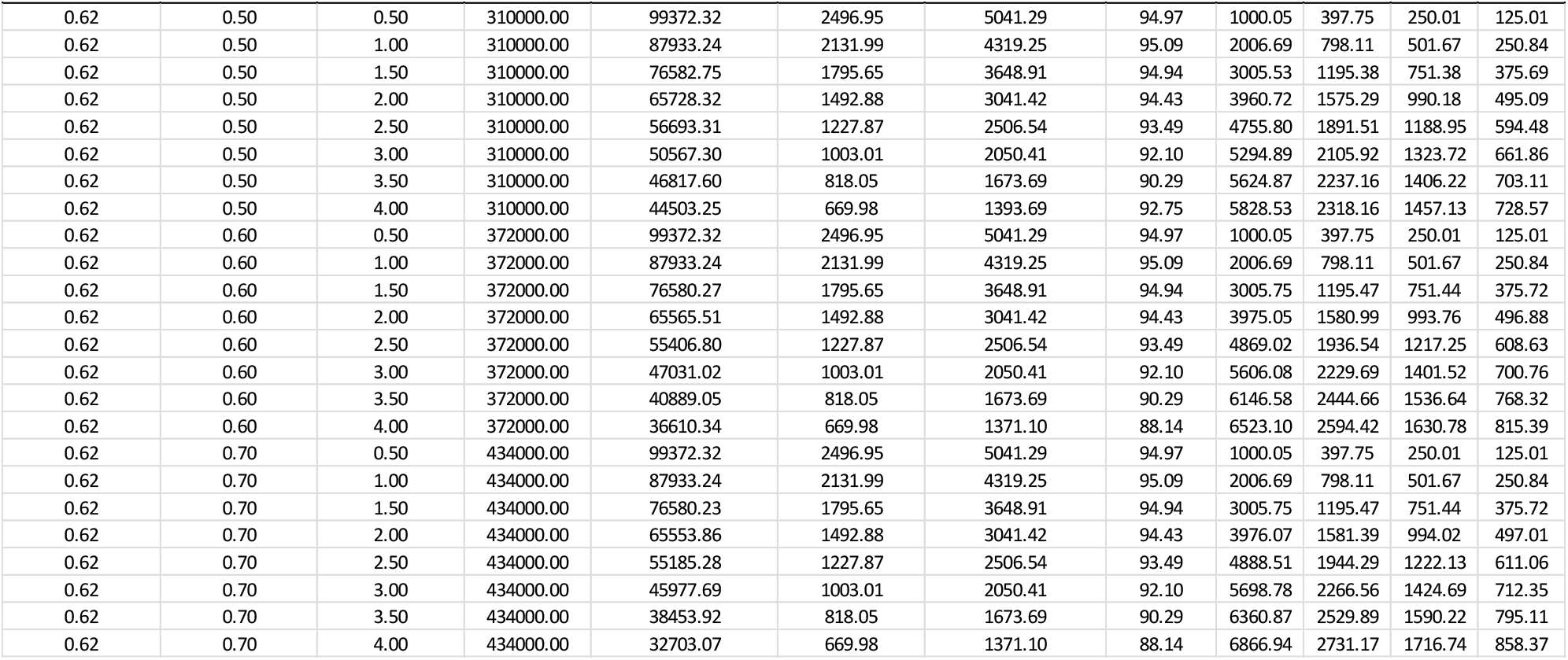
Effect of different vaccination scenarios in relevant indicators of the local evolution of pandemic COVID-19. Indicators are calculated for different vaccination scenarios, including the day of the epidemic peak, the number of new infection cases at the epidemic peak, the cumulative number of symptomatic infections after 150 days of the local pandemic onset, the maximum bed occupancy, and the number of fatalities at different case fatality rates.

## References

1. Oliver SE, Gargano JW, Marin M, Wallace M, Curran KG, Chamberland M, et al. The Advisory Committee on Immunization Practices’ Interim Recommendation for Use of Moderna COVID-19 Vaccine — United States, December 2020. MMWR Morb Mortal Wkly Rep. 2021;69: 1653–1656. doi:10.15585/mmwr.mm695152e1

2. Tanne JH. Covid-19: FDA approves Moderna vaccine as US starts vaccinating health workers [Internet]. The BMJ. BMJ Publishing Group; 2020. doi:10.1136/bmj.m4924

3. Tanne JH. Covid-19: FDA panel votes to approve Pfizer BioNTech vaccine. BMJ. NLM (Medline); 2020;371: m4799. doi:10.1136/bmj.m4799

4. Jackson LA, Anderson EJ, Rouphael NG, Roberts PC, Makhene M, Coler RN, et al. An mRNA Vaccine against SARS-CoV-2 — Preliminary Report. N Engl J Med. Massachusetts Medical Society; 2020;383: 1920–1931. doi:10.1056/nejmoa2022483

5. Polack FP, Thomas SJ, Kitchin N, Absalon J, Gurtman A, Lockhart S, et al. Safety and Efficacy of the BNT162b2 mRNA Covid-19 Vaccine. N Engl J Med. Massachusetts Medical Society; 2020;383: 2603–2615. doi:10.1056/NEJMoa2034577

6. Baden LR, El Sahly HM, Essink B, Kotloff K, Frey S, Novak R, et al. Efficacy and Safety of the mRNA-1273 SARS-CoV-2 Vaccine. N Engl J Med. Massachusetts Medical Society; 2020; NEJMoa2035389. doi:10.1056/NEJMoa2035389

7. Mahase E. Covid-19: What do we know about the late stage vaccine candidates? BMJ. NLM (Medline); 2020;371: m4576. doi:10.1136/bmj.m4576

8. Anderson RM, Vegvari C, Truscott J, Collyer BS. Challenges in creating herd immunity to SARS-CoV-2 infection by mass vaccination [Internet]. The Lancet. Lancet Publishing Group; 2020. pp. 1614–1616. doi:10.1016/S0140-6736(20)32318-7

9. Mello MM, Silverman RD, Omer SB. Ensuring Uptake of Vaccines against SARS- CoV-2. N Engl J Med. Massachusetts Medical Society; 2020;383: 1296–1299. doi:10.1056/nejmp2020926

10. Mahase E. Covid-19: Moderna vaccine is nearly 95% effective, trial involving high risk and elderly people shows. doi:10.1136/bmj.m4347

11. Bartsch S, O’Shea K, … MF-A journal of, 2020 undefined. Vaccine efficacy needed for a COVID-19 coronavirus vaccine to prevent or stop an epidemic as the sole intervention. Elsevier. Available: https://www.sciencedirect.com/science/article/pii/S0749379720302841?casa_token=eM6gGpeNi3UAAAAA:dCIo_2EUT_ooEtI1RcTAccQvfV3OoUBG916hWuhVqeeOLPNTBChNy8N00XIhBal0zaxFRVfQ1Dg

12. Acuña-Zegarra MA, Díaz-Infante S, Baca-Carrasco D, Liceaga DO. COVID-19 optimal vaccination policies: a modeling study on efficacy, natural and vaccine-induced immunity responses. medRxiv. Cold Spring Harbor Laboratory Press; 2020; 2020.11.19.20235176. doi:10.1101/2020.11.19.20235176

13. Alvarez MM, Gonzalez-Gonzalez E, Santiago GT. Modeling COVID-19 epidemics in an Excel spreadsheet: Democratizing the access to first-hand accurate predictions of epidemic outbreaks. medRxiv. Cold Spring Harbor Laboratory Press; 2020; 2020.03.23.20041590. doi:10.1101/2020.03.23.20041590

14. Alvarez MM, Trujillo-de Santiago G. A novel demographic-based model shows that intensive testing and social distancing are concurrently required to extinguish COVID-19 progression in densely populated urban areas. medRxiv. Cold Spring Harbor Laboratory Press; 2020; 2020.06.23.20138743. doi:10.1101/2020.06.23.20138743

15. Robinson E, Jones A, Lesser I, Daly M. International estimates of intended uptake and refusal of COVID-19 vaccines: A rapid systematic review and meta-analysis of large nationally representative samples. medRxiv. Cold Spring Harbor Laboratory Press; 2020; 2020.12.01.20241729. doi:10.1101/2020.12.01.20241729

16. Hasell, Joe; Mathieu, Edouard; Beltekian, Diana; Macdonald, Bobbie; Giattino, Charlie; Ortiz-Ospina, Esteban; Roser, Max; Ritchie H. A cross-country database of COVID-19 testing. Sci data. 2020;7: 1–7. Available: https://www.nature.com/articles/s41597-020-00688-8

17. Bao L, Deng W, Gao H, Xiao C, Liu J, Xue J, et al. Reinfection could not occur in SARS-CoV-2 infected rhesus macaques. bioRxiv. Cold Spring Harbor Laboratory; 2020; 2020.03.13.990226. doi:10.1101/2020.03.13.990226

18. Prompetchara E, Ketloy C, Palaga T. Allergy and Immunology Immune responses in COVID-19 and potential vaccines: Lessons learned from SARS and MERS epidemic. doi:10.12932/AP-200220-0772

19. Liu W, Fontanet A, Zhang P, Zhan L, Xin Z, Baril L, et al. Two-Year Prospective Study of the Humoral Immune Response of Patients with Severe Acute Respiratory Syndrome. J Infect Dis. Oxford University Press (OUP); 2006;193: 792–795. doi:10.1086/500469

20. Subramanian R, He Q, Pascual M. D R A F T Quantifying Asymptomatic Infection and Transmission of COVID-19 in New York City using Observed Cases, Serology and Testing Capacity. medRxiv. Cold Spring Harbor Laboratory Press; 2020; 2020.10.16.20214049. doi:10.1073/

21. Lan L, Xu D, Ye G, Xia C, Wang S, Li Y, et al. Positive RT-PCR Test Results in Patients Recovered From COVID-19. JAMA. 2020; doi:10.1001/jama.2020.2783

22. Bi Q, Wu Y, Mei S, Ye C, Zou X, Zhang Z, et al. Epidemiology and Transmission of COVID-19 in Shenzhen China: Analysis of 391 cases and 1,286 of their close contacts. medRxiv. Cold Spring Harbor Laboratory Press; 2020; 2020.03.03.20028423. doi:10.1101/2020.03.03.20028423

23. Wölfel R, Corman VM, Guggemos W, Seilmaier M, Zange S, Müller MA, et al. Virological assessment of hospitalized patients with COVID-2019. Nature. Nature Research; 2020;581: 465–469. doi:10.1038/s41586-020-2196-x

24. Zou L, Ruan F, Huang M, Liang L, Huang H, Hong Z, et al. SARS-CoV-2 Viral Load in Upper Respiratory Specimens of Infected Patients. N Engl J Med. Massachusetts Medical Society; 2020;382: 1177–1179. doi:10.1056/NEJMc2001737

25. Bai Y, Yao L, Wei T, Tian F, Jin D-Y, Chen L, et al. Presumed Asymptomatic Carrier Transmission of COVID-19. JAMA. 2020; doi:10.1001/jama.2020.2565

26. MacIntyre CR. Global spread of COVID-19 and pandemic potential. Glob Biosecurity. University of New South Wales; 2020;1. doi:10.31646/gbio.55

27. Home - Johns Hopkins Coronavirus Resource Center [Internet]. [cited 10 Sep 2020]. Available: https://coronavirus.jhu.edu/

28. Lewnard JA, Liu VX, Jackson ML, Schmidt MA, Jewell BL, Flores JP, et al. Incidence, clinical outcomes, and transmission dynamics of severe coronavirus disease 2019 in California and Washington: Prospective cohort study. BMJ. BMJ Publishing Group; 2020;369. doi:10.1136/bmj.m1923

29. Guan W, Ni Z, Hu Y, Liang W, Ou C, He J, et al. Clinical Characteristics of Coronavirus Disease 2019 in China. N Engl J Med. Massachussetts Medical Society; 2020;382: 1708–1720. doi:10.1056/NEJMoa2002032

30. Pan L, Mu M, Yang P, Sun Y, Wang R, Yan J, et al. Clinical characteristics of COVID-19 patients with digestive symptoms in Hubei, China: A descriptive, cross-sectional, multicenter study. Am J Gastroenterol. Wolters Kluwer Health; 2020;115: 766–773. doi:10.14309/ajg.0000000000000620

31. Rosenbaum L. Facing Covid-19 in Italy - Ethics, Logistics, and Therapeutics on the Epidemic’s Front Line. N Engl J Med. NLM (Medline); 2020;382: 1873–1875. doi:10.1056/NEJMp2005492

