## Supplemental Information for "Modeling the effect of vaccination strategies in an Excel spreadsheet: The rate of vaccination, and not only the vaccination coverage, is a determinant for containing COVID-19 in urban areas"

### Supplementary information

Mario Moisés Alvarez[1,2*], Sergio Bravo-González[1,2], and Grissel Trujillo-de Santiago[1,3]

[1] Centro de Biotecnología-FEMSA, Tecnologico de Monterrey, Monterrey 64849, NL, México

[2] Departamento de Bioingeniería, Escuela de Ingeniería y Ciencias, Tecnologico de Monterrey, Monterrey 64849, NL, México

[3] Departamento de Ingeniería Mecatrónica y Eléctrica, Escuela de Ingeniería y Ciencias, Tecnologico de Monterrey, Monterrey 64849, NL, México

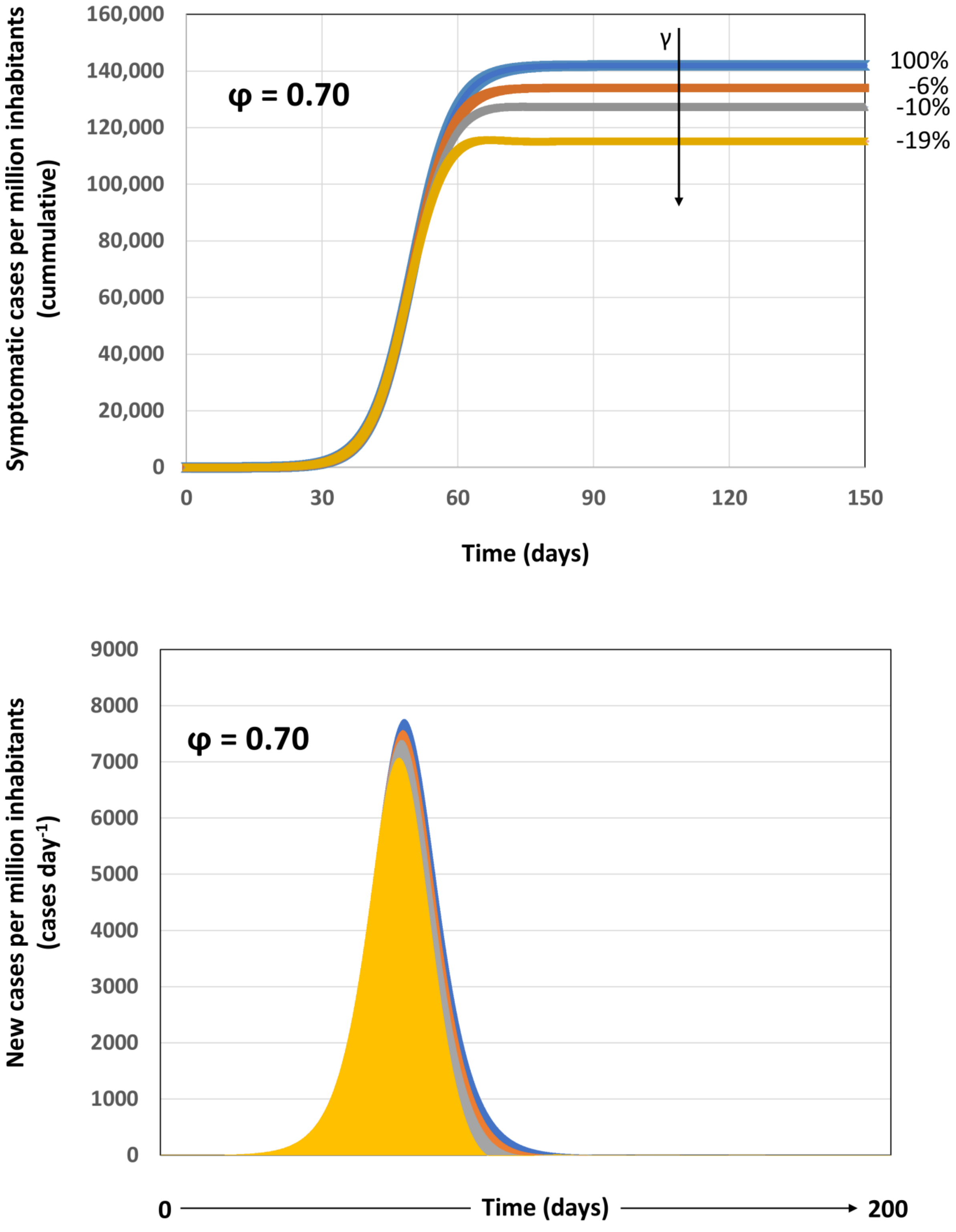


**Figure S1.** S**cenarios of pandemic evolution during the vaccination of 70% of the population at different vaccination rates under conditions of modest social distancing (i.e., 25%).** (A) The cumulative number of infections, (B) the number of new infections per day and the maximum bed occupancy (inset) are presented for vaccination scenarios in which the vaccine coverage is kept constant at 70% $P_o$ ($\varphi$=0.70), the effectiveness of the social distancing measures is 20% ($\sigma$=0.20), a basal level of testing is established ($\alpha$=0.15), and different vaccination rates are imposed, such that: the entire population could be vaccinated within a year (vaccination at $\gamma$=0.0027 $P_o$ day$^{-1}$; orange curve), within six months (vaccination at $\gamma$=0.0054 $P_o$ day$^{-1}$; grey curve), or within three months (vaccination at $\gamma$=0.0108 $P_o$ day$^{-1}$; yellow curve). A reference scenario without vaccination is included (blue line). Numbers indicate the percentage of reduction of symptomatic cases that results from the application of each vaccination rate with respect to the reference case.

**Table S1.** Specific infection rates ($\mu_o$) and the associated doubling times ($t_d$) for COVID-19 infection in different geographic regions. Note that the intrinsic rate of infection ranges between 0.30 and 0.37 for territories with population densities between 4,000 and 7,000 hab km$^2$.

| **Territory** | **Population density** (hab km$^2$) | **μ** (day$^{-1}$) | **$t_d$** (day) |
|---|---|---|---|
| Sao Paulo; Brazil | ~7,000 | 0.308 | 2.250 |
| Mexico City; México | 6,000 | 0.329 | 2.107 |
| Toronto; Canada | 4,149 | 0.330 | 2.100 |
| Lisbon; Portugal | 5,476 | 0.341 | 2.032 |
| Madrid; Spain | 5,418 | 0.358 | 1.937 |
| London; England | 5,701 | 0.362 | 1.915 |
| Amsterdam; Netherlands | 4,439 | 0.424 | 1.635 |
| NYC; USA | 10,194 | 0.666 | 1.040 |

**Table S2.** Effect of different vaccination scenarios in relevant indicators of the local evolution of pandemic COVID-19. Indicators are calculated for different vaccination scenarios, including the day of the epidemic peak, the number of new infection cases at the epidemic peak, the cumulative number of symptomatic infections after 150 days of the local pandemic onset, the maximum bed occupancy, and the number of fatalities at different case fatality rates.

| Vaccination parameters | | | | Local pandemic indicators | | | | Case fatality rates | | | |
|---|---|---|---|---|---|---|---|---|---|---|---|
| η: Vaccine Efficacy | ϕ: Vaccination Coverage | vaccination rate ($\gamma$ Po year$^{-1}$) | vaccine equivalents | max_symptomatic | max_new cases | max_bed occupancy | peak @ day | 0.09 | 0.04 | 0.02 | 0.01 |
| 0.95 | 0.30 | 0.00 | 0.00 | 110736.53 | 2885.67 | 5804.24 | 94.66 | 0.00 | 0.00 | 0.00 | 0.00 |
| 0.95 | 0.30 | 0.50 | 285000.00 | 93284.68 | 2299.44 | 4651.22 | 95.06 | 1535.76 | 610.81 | 383.94 | 191.97 |
| 0.95 | 0.30 | 1.00 | 285000.00 | 75870.70 | 1775.05 | 3607.71 | 94.92 | 3068.19 | 1220.30 | 767.05 | 383.52 |
| 0.95 | 0.30 | 1.50 | 285000.00 | 60820.34 | 1329.98 | 2712.98 | 93.92 | 4392.62 | 1747.07 | 1098.16 | 549.08 |
| 0.95 | 0.30 | 2.00 | 285000.00 | 52753.41 | 976.94 | 1997.40 | 91.89 | 5102.51 | 2029.41 | 1275.63 | 637.81 |
| 0.95 | 0.30 | 2.50 | 285000.00 | 49332.18 | 740.92 | 1535.01 | 94.96 | 5403.58 | 2149.15 | 1350.90 | 675.45 |
| 0.95 | 0.30 | 3.00 | 285000.00 | 47699.85 | 644.99 | 1340.75 | 100.07 | 5547.23 | 2206.28 | 1386.81 | 693.40 |
| 0.95 | 0.30 | 3.50 | 285000.00 | 46784.89 | 598.89 | 1246.27 | 104.32 | 5627.74 | 2238.31 | 1406.94 | 703.47 |
| 0.95 | 0.30 | 4.00 | 285000.00 | 46204.29 | 573.15 | 1193.17 | 107.57 | 5678.84 | 2258.63 | 1419.71 | 709.85 |
| 0.95 | 0.40 | 0.50 | 380000.00 | 93284.68 | 2299.44 | 4651.22 | 95.06 | 1535.76 | 610.81 | 383.94 | 191.97 |
| 0.95 | 0.40 | 1.00 | 380000.00 | 75856.01 | 1775.05 | 3607.71 | 94.92 | 3069.49 | 1220.82 | 767.37 | 383.69 |
| 0.95 | 0.40 | 1.50 | 380000.00 | 59319.86 | 1329.98 | 2712.98 | 93.92 | 4524.67 | 1799.58 | 1131.17 | 565.58 |
| 0.95 | 0.40 | 2.00 | 380000.00 | 45856.77 | 976.94 | 1997.40 | 91.89 | 5709.42 | 2270.79 | 1427.35 | 713.68 |
| 0.95 | 0.40 | 2.50 | 380000.00 | 37260.87 | 716.28 | 1465.82 | 88.90 | 6465.86 | 2571.65 | 1616.46 | 808.23 |
| 0.95 | 0.40 | 3.00 | 380000.00 | 32212.76 | 534.43 | 1093.37 | 85.33 | 6910.09 | 2748.33 | 1727.52 | 863.76 |
| 0.95 | 0.40 | 3.50 | 380000.00 | 29088.93 | 411.00 | 842.92 | 86.74 | 7184.99 | 2857.67 | 1796.25 | 898.12 |
| 0.95 | 0.40 | 4.00 | 380000.00 | 26996.63 | 327.16 | 680.96 | 86.01 | 7369.11 | 2930.90 | 1842.28 | 921.14 |
| 0.95 | 0.50 | 0.50 | 475000.00 | 93284.68 | 2299.44 | 4651.22 | 95.06 | 1535.76 | 610.81 | 383.94 | 191.97 |
| 0.95 | 0.50 | 1.00 | 475000.00 | 75855.97 | 1775.05 | 3607.71 | 94.92 | 3069.49 | 1220.82 | 767.37 | 383.69 |
| 0.95 | 0.50 | 1.50 | 475000.00 | 59253.45 | 1329.98 | 2712.98 | 93.92 | 4530.51 | 1801.91 | 1132.63 | 566.31 |
| 0.95 | 0.50 | 2.00 | 475000.00 | 44678.68 | 976.94 | 1997.40 | 91.89 | 5813.09 | 2312.02 | 1453.27 | 726.64 |
| 0.95 | 0.50 | 2.50 | 475000.00 | 33502.37 | 716.28 | 1465.82 | 88.90 | 6796.61 | 2703.20 | 1699.15 | 849.58 |
| 0.95 | 0.50 | 3.00 | 475000.00 | 25909.36 | 534.43 | 1093.37 | 85.33 | 7464.79 | 2968.95 | 1866.20 | 933.10 |
| 0.95 | 0.50 | 3.50 | 475000.00 | 20899.80 | 411.00 | 839.88 | 81.64 | 7905.63 | 3144.29 | 1976.41 | 988.20 |
| 0.95 | 0.50 | 4.00 | 475000.00 | 17504.14 | 327.16 | 667.39 | 78.13 | 8204.45 | 3263.13 | 2051.11 | 1025.56 |
| 0.95 | 0.60 | 0.50 | 570000.00 | 93284.68 | 2299.44 | 4651.22 | 95.06 | 1535.76 | 610.81 | 383.94 | 191.97 |
| 0.95 | 0.60 | 1.00 | 570000.00 | 75855.97 | 1775.05 | 3607.71 | 94.92 | 3069.49 | 1220.82 | 767.37 | 383.69 |
| 0.95 | 0.60 | 1.50 | 570000.00 | 59251.93 | 1329.98 | 2712.98 | 93.92 | 4530.64 | 1801.96 | 1132.66 | 566.33 |
| 0.95 | 0.60 | 2.00 | 570000.00 | 44582.17 | 976.94 | 1997.40 | 91.89 | 5821.58 | 2315.40 | 1455.40 | 727.70 |
| 0.95 | 0.60 | 2.50 | 570000.00 | 32839.29 | 716.28 | 1465.82 | 88.90 | 6854.96 | 2726.40 | 1713.74 | 856.87 |
| 0.95 | 0.60 | 3.00 | 570000.00 | 24346.79 | 534.43 | 1093.37 | 85.33 | 7602.30 | 3023.64 | 1900.57 | 950.29 |
| 0.95 | 0.60 | 3.50 | 570000.00 | 18561.72 | 411.00 | 839.88 | 81.64 | 8111.38 | 3226.12 | 2027.85 | 1013.92 |
| 0.95 | 0.60 | 4.00 | 570000.00 | 14652.20 | 327.16 | 667.39 | 78.13 | 8455.42 | 3362.95 | 2113.86 | 1056.93 |
| 0.95 | 0.70 | 0.50 | 665000.00 | 93284.68 | 2299.44 | 4651.22 | 95.06 | 1535.76 | 610.81 | 383.94 | 191.97 |
| 0.95 | 0.70 | 1.00 | 665000.00 | 75855.97 | 1775.05 | 3607.71 | 94.92 | 3069.49 | 1220.82 | 767.37 | 383.69 |
| 0.95 | 0.70 | 1.50 | 665000.00 | 59251.89 | 1329.98 | 2712.98 | 93.92 | 4530.65 | 1801.96 | 1132.66 | 566.33 |
| 0.95 | 0.70 | 2.00 | 665000.00 | 44577.76 | 976.94 | 1997.40 | 91.89 | 5821.97 | 2315.56 | 1455.49 | 727.75 |
| 0.95 | 0.70 | 2.50 | 665000.00 | 32767.47 | 716.28 | 1465.82 | 88.90 | 6861.28 | 2728.92 | 1715.32 | 857.66 |
| 0.95 | 0.70 | 3.00 | 665000.00 | 24070.20 | 534.43 | 1093.37 | 85.33 | 7626.64 | 3033.32 | 1906.66 | 953.33 |
| 0.95 | 0.70 | 3.50 | 665000.00 | 18037.42 | 411.00 | 839.88 | 81.64 | 8157.52 | 3244.47 | 2039.38 | 1019.69 |
| 0.95 | 0.70 | 4.00 | 665000.00 | 13936.55 | 327.16 | 667.39 | 78.13 | 8518.40 | 3388.00 | 2129.60 | 1064.80 |
| 0.90 | 0.30 | 0.50 | 270000.00 | 94207.71 | 2328.90 | 4709.52 | 95.05 | 1454.54 | 578.51 | 363.63 | 181.82 |
| 0.90 | 0.30 | 1.00 | 270000.00 | 77684.00 | 1826.81 | 3711.21 | 94.97 | 2908.62 | 1156.84 | 727.16 | 363.58 |
| 0.90 | 0.30 | 1.50 | 270000.00 | 63345.18 | 1394.35 | 2842.89 | 94.15 | 4170.44 | 1658.70 | 1042.61 | 521.30 |
| 0.90 | 0.30 | 2.00 | 270000.00 | 55747.12 | 1043.37 | 2132.43 | 92.41 | 4839.07 | 1924.63 | 1209.77 | 604.88 |
| 0.90 | 0.30 | 2.50 | 270000.00 | 52618.16 | 811.28 | 1679.55 | 95.75 | 5114.42 | 2034.14 | 1278.60 | 639.30 |
| 0.90 | 0.30 | 3.00 | 270000.00 | 51168.80 | 719.24 | 1493.50 | 100.61 | 5241.96 | 2084.87 | 1310.49 | 655.24 |
| 0.90 | 0.30 | 3.50 | 270000.00 | 50376.47 | 675.32 | 1403.68 | 104.44 | 5311.68 | 2112.60 | 1327.92 | 663.96 |
| 0.90 | 0.30 | 4.00 | 270000.00 | 49884.00 | 650.89 | 1353.42 | 107.32 | 5355.02 | 2129.84 | 1338.76 | 669.38 |
| 0.90 | 0.40 | 0.50 | 360000.00 | 94207.71 | 2328.90 | 4709.52 | 95.05 | 1454.54 | 578.51 | 363.63 | 181.82 |
| 0.90 | 0.40 | 1.00 | 360000.00 | 77669.13 | 1826.81 | 3711.21 | 94.97 | 2909.93 | 1157.36 | 727.48 | 363.74 |
| 0.90 | 0.40 | 1.50 | 360000.00 | 61846.48 | 1394.35 | 2842.89 | 94.15 | 4302.32 | 1711.15 | 1075.58 | 537.79 |
| 0.90 | 0.40 | 2.00 | 360000.00 | 48826.31 | 1043.37 | 2132.43 | 92.41 | 5448.10 | 2166.86 | 1362.02 | 681.01 |
| 0.90 | 0.40 | 2.50 | 360000.00 | 40549.04 | 776.48 | 1588.83 | 89.76 | 6176.50 | 2456.56 | 1544.12 | 772.06 |
| 0.90 | 0.40 | 3.00 | 360000.00 | 35780.66 | 584.55 | 1196.15 | 86.49 | 6596.12 | 2623.46 | 1649.03 | 824.51 |
| 0.90 | 0.40 | 3.50 | 360000.00 | 32893.12 | 450.96 | 932.62 | 88.52 | 6850.22 | 2724.52 | 1712.55 | 856.28 |
| 0.90 | 0.40 | 4.00 | 360000.00 | 30993.56 | 369.73 | 770.27 | 89.10 | 7017.38 | 2791.00 | 1754.35 | 877.17 |
| 0.90 | 0.50 | 0.50 | 450000.00 | 94207.71 | 2328.90 | 4709.52 | 95.05 | 1454.54 | 578.51 | 363.63 | 181.82 |
| 0.90 | 0.50 | 1.00 | 450000.00 | 77669.08 | 1826.81 | 3711.21 | 94.97 | 2909.93 | 1157.36 | 727.48 | 363.74 |
| 0.90 | 0.50 | 1.50 | 450000.00 | 61776.59 | 1394.35 | 2842.89 | 94.15 | 4308.47 | 1713.60 | 1077.12 | 538.56 |
| 0.90 | 0.50 | 2.00 | 450000.00 | 47568.23 | 1043.37 | 2132.43 | 92.41 | 5558.81 | 2210.89 | 1389.70 | 694.85 |
| 0.90 | 0.50 | 2.50 | 450000.00 | 36449.84 | 776.48 | 1588.83 | 89.76 | 6537.23 | 2600.03 | 1634.31 | 817.15 |
| 0.90 | 0.50 | 3.00 | 450000.00 | 28808.50 | 584.55 | 1196.15 | 86.49 | 7209.67 | 2867.48 | 1802.42 | 901.21 |
| 0.90 | 0.50 | 3.50 | 450000.00 | 23739.71 | 450.96 | 922.02 | 82.99 | 7655.72 | 3044.89 | 1913.93 | 956.97 |
| 0.90 | 0.50 | 4.00 | 450000.00 | 20286.17 | 358.64 | 732.19 | 79.57 | 7959.63 | 3165.76 | 1989.91 | 994.95 |

**Table S2.** (continuation) Effect of different vaccination scenarios in relevant indicators of the local evolution of pandemic COVID-19.

| Vaccination parameters | | | | Local pandemic indicators | | | | Case fatality rates | | | |
|---|---|---|---|---|---|---|---|---|---|---|---|
| η: Vaccine Efficacy | ϕ: Vaccination Coverage | vaccination rate ($\gamma$ Po year$^{-1}$) | vaccine equivalents | max_symptomatic | max_new cases | max_bed occupancy | peak @ day | 0.09 | 0.04 | 0.02 | 0.01 |
| 0.90 | 0.60 | 0.50 | 540000.00 | 94207.71 | 2328.90 | 4709.52 | 95.05 | 1454.54 | 578.51 | 363.63 | 181.82 |
| 0.90 | 0.60 | 1.00 | 540000.00 | 77669.08 | 1826.81 | 3711.21 | 94.97 | 2909.93 | 1157.36 | 727.48 | 363.74 |
| 0.90 | 0.60 | 1.50 | 540000.00 | 61774.93 | 1394.35 | 2842.89 | 94.15 | 4308.62 | 1713.66 | 1077.16 | 538.58 |
| 0.90 | 0.60 | 2.00 | 540000.00 | 47458.01 | 1043.37 | 2132.43 | 92.41 | 5568.51 | 2214.75 | 1392.13 | 696.06 |
| 0.90 | 0.60 | 2.50 | 540000.00 | 35666.55 | 776.48 | 1588.83 | 89.76 | 6606.16 | 2627.45 | 1651.54 | 825.77 |
| 0.90 | 0.60 | 3.00 | 540000.00 | 26898.28 | 584.55 | 1196.15 | 86.49 | 7377.77 | 2934.34 | 1844.44 | 922.22 |
| 0.90 | 0.60 | 3.50 | 540000.00 | 20798.17 | 450.96 | 922.02 | 82.99 | 7914.58 | 3147.84 | 1978.64 | 989.32 |
| 0.90 | 0.60 | 4.00 | 540000.00 | 16609.98 | 358.64 | 732.19 | 79.57 | 8283.14 | 3294.43 | 2070.78 | 1035.39 |
| 0.90 | 0.70 | 0.50 | 630000.00 | 94207.71 | 2328.90 | 4709.52 | 95.05 | 1454.54 | 578.51 | 363.63 | 181.82 |
| 0.90 | 0.70 | 1.00 | 630000.00 | 77669.08 | 1826.81 | 3711.21 | 94.97 | 2909.93 | 1157.36 | 727.48 | 363.74 |
| 0.90 | 0.70 | 1.50 | 630000.00 | 61774.89 | 1394.35 | 2842.89 | 94.15 | 4308.62 | 1713.66 | 1077.16 | 538.58 |
| 0.90 | 0.70 | 2.00 | 630000.00 | 47452.65 | 1043.37 | 2132.43 | 92.41 | 5568.98 | 2214.94 | 1392.25 | 696.12 |
| 0.90 | 0.70 | 2.50 | 630000.00 | 35574.16 | 776.48 | 1588.83 | 89.76 | 6614.29 | 2630.68 | 1653.57 | 826.79 |
| 0.90 | 0.70 | 3.00 | 630000.00 | 26529.61 | 584.55 | 1196.15 | 86.49 | 7410.21 | 2947.24 | 1852.55 | 926.28 |
| 0.90 | 0.70 | 3.50 | 630000.00 | 20079.36 | 450.96 | 922.02 | 82.99 | 7977.83 | 3173.00 | 1994.46 | 997.23 |
| 0.90 | 0.70 | 4.00 | 630000.00 | 15608.76 | 358.64 | 732.19 | 79.57 | 8371.24 | 3329.47 | 2092.81 | 1046.41 |
| 0.70 | 0.30 | 0.50 | 210000.00 | 97897.87 | 2448.43 | 4945.61 | 95.00 | 1129.80 | 449.35 | 282.45 | 141.23 |
| 0.70 | 0.30 | 1.00 | 210000.00 | 85001.84 | 2042.25 | 4140.86 | 95.08 | 2264.65 | 900.71 | 566.16 | 283.08 |
| 0.70 | 0.30 | 1.50 | 210000.00 | 73707.24 | 1674.21 | 3405.71 | 94.79 | 3258.58 | 1296.02 | 814.64 | 407.32 |
| 0.70 | 0.30 | 2.00 | 210000.00 | 67988.32 | 1351.18 | 2757.03 | 94.00 | 3761.84 | 1496.19 | 940.46 | 470.23 |
| 0.70 | 0.30 | 2.50 | 210000.00 | 65871.32 | 1147.37 | 2366.24 | 97.40 | 3948.14 | 1570.28 | 987.03 | 493.52 |
| 0.70 | 0.30 | 3.00 | 210000.00 | 64986.34 | 1072.12 | 2214.70 | 100.72 | 4026.02 | 1601.26 | 1006.50 | 503.25 |
| 0.70 | 0.30 | 3.50 | 210000.00 | 64542.07 | 1036.88 | 2143.58 | 103.08 | 4065.11 | 1616.81 | 1016.28 | 508.14 |
| 0.70 | 0.30 | 4.00 | 210000.00 | 64284.98 | 1017.70 | 2104.38 | 104.82 | 4087.74 | 1625.80 | 1021.93 | 510.97 |
| 0.70 | 0.40 | 0.50 | 280000.00 | 97897.87 | 2448.43 | 4945.61 | 95.00 | 1129.80 | 449.35 | 282.45 | 141.23 |
| 0.70 | 0.40 | 1.00 | 280000.00 | 84986.99 | 2042.25 | 4140.86 | 95.08 | 2265.96 | 901.23 | 566.49 | 283.24 |
| 0.70 | 0.40 | 1.50 | 280000.00 | 72336.78 | 1674.21 | 3405.71 | 94.79 | 3379.18 | 1343.99 | 844.79 | 422.40 |
| 0.70 | 0.40 | 2.00 | 280000.00 | 61658.26 | 1351.18 | 2755.79 | 94.00 | 4318.89 | 1717.74 | 1079.72 | 539.86 |
| 0.70 | 0.40 | 2.50 | 280000.00 | 55130.50 | 1078.16 | 2203.08 | 92.65 | 4893.33 | 1946.21 | 1223.33 | 611.67 |
| 0.70 | 0.40 | 3.00 | 280000.00 | 51747.01 | 856.45 | 1769.11 | 93.90 | 5191.08 | 2064.63 | 1297.77 | 648.88 |
| 0.70 | 0.40 | 3.50 | 280000.00 | 49928.86 | 731.94 | 1519.45 | 96.14 | 5351.07 | 2128.27 | 1337.77 | 668.88 |
| 0.70 | 0.40 | 4.00 | 280000.00 | 48855.26 | 669.08 | 1390.35 | 100.28 | 5445.55 | 2165.84 | 1361.39 | 680.69 |
| 0.70 | 0.50 | 0.50 | 350000.00 | 97897.87 | 2448.43 | 4945.61 | 95.00 | 1129.80 | 449.35 | 282.45 | 141.23 |
| 0.70 | 0.50 | 1.00 | 350000.00 | 84986.95 | 2042.25 | 4140.86 | 95.08 | 2265.96 | 901.24 | 566.49 | 283.25 |
| 0.70 | 0.50 | 1.50 | 350000.00 | 72259.67 | 1674.21 | 3405.71 | 94.79 | 3385.96 | 1346.69 | 846.49 | 423.25 |
| 0.70 | 0.50 | 2.00 | 350000.00 | 60252.91 | 1351.18 | 2755.79 | 94.00 | 4442.56 | 1766.93 | 1110.64 | 555.32 |
| 0.70 | 0.50 | 2.50 | 350000.00 | 50351.96 | 1078.16 | 2203.08 | 92.65 | 5313.84 | 2113.46 | 1328.46 | 664.23 |
| 0.70 | 0.50 | 3.00 | 350000.00 | 43534.03 | 856.45 | 1752.00 | 90.73 | 5913.82 | 2352.09 | 1478.45 | 739.23 |
| 0.70 | 0.50 | 3.50 | 350000.00 | 39194.02 | 682.82 | 1397.39 | 88.36 | 6295.74 | 2503.99 | 1573.94 | 786.97 |
| 0.70 | 0.50 | 4.00 | 350000.00 | 36384.98 | 550.46 | 1130.12 | 90.65 | 6542.94 | 2602.30 | 1635.73 | 817.87 |
| 0.70 | 0.60 | 0.50 | 420000.00 | 97897.87 | 2448.43 | 4945.61 | 95.00 | 1129.80 | 449.35 | 282.45 | 141.23 |
| 0.70 | 0.60 | 1.00 | 420000.00 | 84986.95 | 2042.25 | 4140.86 | 95.08 | 2265.96 | 901.24 | 566.49 | 283.25 |
| 0.70 | 0.60 | 1.50 | 420000.00 | 72257.41 | 1674.21 | 3405.71 | 94.79 | 3386.16 | 1346.77 | 846.54 | 423.27 |
| 0.70 | 0.60 | 2.00 | 420000.00 | 60098.36 | 1351.18 | 2755.79 | 94.00 | 4456.16 | 1772.34 | 1114.04 | 557.02 |
| 0.70 | 0.60 | 2.50 | 420000.00 | 49147.06 | 1078.16 | 2203.08 | 92.65 | 5419.87 | 2155.63 | 1354.97 | 677.48 |
| 0.70 | 0.60 | 3.00 | 420000.00 | 40280.86 | 856.45 | 1752.00 | 90.73 | 6200.10 | 2465.95 | 1550.02 | 775.01 |
| 0.70 | 0.60 | 3.50 | 420000.00 | 33772.57 | 682.82 | 1397.39 | 88.36 | 6772.83 | 2693.74 | 1693.21 | 846.60 |
| 0.70 | 0.60 | 4.00 | 420000.00 | 29160.43 | 550.46 | 1126.25 | 85.72 | 7178.70 | 2855.16 | 1794.67 | 897.34 |
| 0.70 | 0.70 | 0.50 | 490000.00 | 97897.87 | 2448.43 | 4945.61 | 95.00 | 1129.80 | 449.35 | 282.45 | 141.23 |
| 0.70 | 0.70 | 1.00 | 490000.00 | 84986.95 | 2042.25 | 4140.86 | 95.08 | 2265.96 | 901.24 | 566.49 | 283.25 |
| 0.70 | 0.70 | 1.50 | 490000.00 | 72257.36 | 1674.21 | 3405.71 | 94.79 | 3386.17 | 1346.77 | 846.54 | 423.27 |
| 0.70 | 0.70 | 2.00 | 490000.00 | 60088.43 | 1351.18 | 2755.79 | 94.00 | 4457.03 | 1772.68 | 1114.26 | 557.13 |
| 0.70 | 0.70 | 2.50 | 490000.00 | 48957.66 | 1078.16 | 2203.08 | 92.65 | 5436.54 | 2162.26 | 1359.14 | 679.57 |
| 0.70 | 0.70 | 3.00 | 490000.00 | 39413.39 | 856.45 | 1752.00 | 90.73 | 6276.44 | 2496.31 | 1569.11 | 784.55 |
| 0.70 | 0.70 | 3.50 | 490000.00 | 31843.05 | 682.82 | 1397.39 | 88.36 | 6942.63 | 2761.27 | 1735.66 | 867.83 |
| 0.70 | 0.70 | 4.00 | 490000.00 | 26157.75 | 550.46 | 1126.25 | 85.72 | 7442.93 | 2960.26 | 1860.73 | 930.37 |
| 0.62 | 0.30 | 0.50 | 186000.00 | 99372.32 | 2496.95 | 5041.29 | 94.97 | 1000.05 | 397.75 | 250.01 | 125.01 |
| 0.62 | 0.30 | 1.00 | 186000.00 | 87947.72 | 2131.99 | 4319.25 | 95.09 | 2005.41 | 797.61 | 501.35 | 250.68 |
| 0.62 | 0.30 | 1.50 | 186000.00 | 77929.84 | 1795.65 | 3648.91 | 94.94 | 2886.99 | 1148.23 | 721.75 | 360.87 |
| 0.62 | 0.30 | 2.00 | 186000.00 | 72939.81 | 1492.88 | 3043.62 | 94.43 | 3326.11 | 1322.89 | 831.53 | 415.76 |
| 0.62 | 0.30 | 2.50 | 186000.00 | 71157.68 | 1304.71 | 2684.74 | 97.51 | 3482.94 | 1385.26 | 870.73 | 435.37 |
| 0.62 | 0.30 | 3.00 | 186000.00 | 70436.73 | 1236.45 | 2547.92 | 100.25 | 3546.38 | 1410.49 | 886.60 | 443.30 |
| 0.62 | 0.30 | 3.50 | 186000.00 | 70084.12 | 1204.74 | 2484.37 | 102.15 | 3577.41 | 1422.83 | 894.35 | 447.18 |
| 0.62 | 0.30 | 4.00 | 186000.00 | 69884.41 | 1187.65 | 2449.53 | 103.56 | 3594.99 | 1429.82 | 898.75 | 449.37 |
| 0.62 | 0.40 | 0.50 | 248000.00 | 99372.32 | 2496.95 | 5041.29 | 94.97 | 1000.05 | 397.75 | 250.01 | 125.01 |
| 0.62 | 0.40 | 1.00 | 248000.00 | 87933.29 | 2131.99 | 4319.25 | 95.09 | 2006.68 | 798.11 | 501.67 | 250.84 |
| 0.62 | 0.40 | 1.50 | 248000.00 | 76659.23 | 1795.65 | 3648.91 | 94.94 | 2998.80 | 1192.71 | 749.70 | 374.85 |
| 0.62 | 0.40 | 2.00 | 248000.00 | 67102.43 | 1492.88 | 3041.42 | 94.43 | 3839.80 | 1527.19 | 959.95 | 479.98 |
| 0.62 | 0.40 | 2.50 | 248000.00 | 61375.25 | 1227.87 | 2506.54 | 93.49 | 4343.79 | 1727.64 | 1085.95 | 542.97 |
| 0.62 | 0.40 | 3.00 | 248000.00 | 58534.91 | 1003.01 | 2082.45 | 95.17 | 4593.74 | 1827.06 | 1148.44 | 574.22 |
| 0.62 | 0.40 | 3.50 | 248000.00 | 57080.00 | 894.99 | 1854.80 | 97.65 | 4721.77 | 1877.98 | 1180.44 | 590.22 |
| 0.62 | 0.40 | 4.00 | 248000.00 | 56257.27 | 838.55 | 1738.23 | 100.95 | 4794.17 | 1906.77 | 1198.54 | 599.27 |

**Table S2.** (continuation) Effect of different vaccination scenarios in relevant indicators of the local evolution of pandemic COVID-19.

| Vaccination parameters | | | | Local pandemic indicators | | | | Case fatality rates | | | |
|---|---|---|---|---|---|---|---|---|---|---|---|
| η: Vaccine Efficacy | ϕ: Vaccination Coverage | vaccination rate (γ Po year$^{-1}$) | vaccine equivalents | max_symptomatic | max_new cases | max_bed occupancy | peak @ day | 0.09 | 0.04 | 0.02 | 0.01 |
| 0.62 | 0.50 | 0.50 | 310000.00 | 99372.32 | 2496.95 | 5041.29 | 94.97 | 1000.05 | 397.75 | 250.01 | 125.01 |
| 0.62 | 0.50 | 1.00 | 310000.00 | 87933.24 | 2131.99 | 4319.25 | 95.09 | 2006.69 | 798.11 | 501.67 | 250.84 |
| 0.62 | 0.50 | 1.50 | 310000.00 | 76582.75 | 1795.65 | 3648.91 | 94.94 | 3005.53 | 1195.38 | 751.38 | 375.69 |
| 0.62 | 0.50 | 2.00 | 310000.00 | 65728.32 | 1492.88 | 3041.42 | 94.43 | 3960.72 | 1575.29 | 990.18 | 495.09 |
| 0.62 | 0.50 | 2.50 | 310000.00 | 56693.31 | 1227.87 | 2506.54 | 93.49 | 4755.80 | 1891.51 | 1188.95 | 594.48 |
| 0.62 | 0.50 | 3.00 | 310000.00 | 50567.30 | 1003.01 | 2050.41 | 92.10 | 5294.89 | 2105.92 | 1323.72 | 661.86 |
| 0.62 | 0.50 | 3.50 | 310000.00 | 46817.60 | 818.05 | 1673.69 | 90.29 | 5624.87 | 2237.16 | 1406.22 | 703.11 |
| 0.62 | 0.50 | 4.00 | 310000.00 | 44503.25 | 669.98 | 1393.69 | 92.75 | 5828.53 | 2318.16 | 1457.13 | 728.57 |
| 0.62 | 0.60 | 0.50 | 372000.00 | 99372.32 | 2496.95 | 5041.29 | 94.97 | 1000.05 | 397.75 | 250.01 | 125.01 |
| 0.62 | 0.60 | 1.00 | 372000.00 | 87933.24 | 2131.99 | 4319.25 | 95.09 | 2006.69 | 798.11 | 501.67 | 250.84 |
| 0.62 | 0.60 | 1.50 | 372000.00 | 76580.27 | 1795.65 | 3648.91 | 94.94 | 3005.75 | 1195.47 | 751.44 | 375.72 |
| 0.62 | 0.60 | 2.00 | 372000.00 | 65565.51 | 1492.88 | 3041.42 | 94.43 | 3975.05 | 1580.99 | 993.76 | 496.88 |
| 0.62 | 0.60 | 2.50 | 372000.00 | 55406.80 | 1227.87 | 2506.54 | 93.49 | 4869.02 | 1936.54 | 1217.25 | 608.63 |
| 0.62 | 0.60 | 3.00 | 372000.00 | 47031.02 | 1003.01 | 2050.41 | 92.10 | 5606.08 | 2229.69 | 1401.52 | 700.76 |
| 0.62 | 0.60 | 3.50 | 372000.00 | 40889.05 | 818.05 | 1673.69 | 90.29 | 6146.58 | 2444.66 | 1536.64 | 768.32 |
| 0.62 | 0.60 | 4.00 | 372000.00 | 36610.34 | 669.98 | 1371.10 | 88.14 | 6523.10 | 2594.42 | 1630.78 | 815.39 |
| 0.62 | 0.70 | 0.50 | 434000.00 | 99372.32 | 2496.95 | 5041.29 | 94.97 | 1000.05 | 397.75 | 250.01 | 125.01 |
| 0.62 | 0.70 | 1.00 | 434000.00 | 87933.24 | 2131.99 | 4319.25 | 95.09 | 2006.69 | 798.11 | 501.67 | 250.84 |
| 0.62 | 0.70 | 1.50 | 434000.00 | 76580.23 | 1795.65 | 3648.91 | 94.94 | 3005.75 | 1195.47 | 751.44 | 375.72 |
| 0.62 | 0.70 | 2.00 | 434000.00 | 65553.86 | 1492.88 | 3041.42 | 94.43 | 3976.07 | 1581.39 | 994.02 | 497.01 |
| 0.62 | 0.70 | 2.50 | 434000.00 | 55185.28 | 1227.87 | 2506.54 | 93.49 | 4888.51 | 1944.29 | 1222.13 | 611.06 |
| 0.62 | 0.70 | 3.00 | 434000.00 | 45977.69 | 1003.01 | 2050.41 | 92.10 | 5698.78 | 2266.56 | 1424.69 | 712.35 |
| 0.62 | 0.70 | 3.50 | 434000.00 | 38453.92 | 818.05 | 1673.69 | 90.29 | 6360.87 | 2529.89 | 1590.22 | 795.11 |
| 0.62 | 0.70 | 4.00 | 434000.00 | 32703.07 | 669.98 | 1371.10 | 88.14 | 6866.94 | 2731.17 | 1716.74 | 858.37 |
